# Old lineages, new threat: the re-emergence of pneumococcal serotype 14 in adults in Catalonia, Spain (2023-2025)

**DOI:** 10.64898/2026.11.16.26367621

**Authors:** Aida González-Díaz, Sonia Broner, Sara Calvo-Silveria, Inmaculada Grau, M Dolores Quesada, Alba Bellés, Eva del Corral, Ester Picó-Plana, Esther Clapés, Belén Viñado, Dàmaris Berbel, M Antonia Llopis, M Ángeles Domínguez, Sara Martí, Jordi Càmara, Pilar Ciruela, Carmen Ardanuy

## Abstract

**Introduction:** The introduction of pneumococcal conjugate vaccines resulted in a decline in invasive pneumococcal disease (IPD). Nevertheless, recent studies report a re-emergence of vaccine-included serotypes.

**Aim:** To describe the recent re-emergence of serotype 14 IPD among adults in Catalonia and to investigate the factors underlying its resurgence.

**Methods:** We analysed two collections of adult IPD isolates: a historical series from Hospital Universitari de Bellvitge (1996–2025) and a multicentre series from seven tertiary hospitals in Catalonia (2023–2025). Isolates were serotyped (Quellung) and genotyped by PFGE/MLST (1996-2022) or Illumina sequencing (2023-2025). Clinical data were retrospectively collected.

**Results:** The historical series showed that, after peaking in 2009, serotype 14 frequency gradually declined, with only 0-4 cases reported annually between 2016 and 2020. However, the number of serotype 14 isolates increased again in 2025, reaching 11.0% of all isolates at HUB and 10.8% across Catalonia. Genomic data showed that this increase involved multiple previously circulating lineages (GPSC6, GPSC18, GPSC16 and GPSC39), with no dominant clone. Analysis of the capsular locus revealed substantial genetic diversity, with no association between capsular variants and either lineage or year of isolation. A substantial proportion of patients (56.3%) had multiple comorbidities and only 18.8% were appropriately vaccinated.

**Discussion:** The re-emergence of serotype 14 was not associated with a novel lineage or capsular variant. Instead, it may reflect a combination of post-pandemic transmission changes, higher-valency paediatric vaccines, and natural fluctuations in serotype circulation. These findings highlight the limits of herd protection and underscore the need to strengthen adult vaccination, particularly among at-risk populations.

## INTRODUCTION

*Streptococcus pneumoniae* remains one of the leading causes of severe infections attributable to vaccine-preventable bacterial pathogens [1]. The introduction of pneumococcal conjugate vaccines (PCVs) into paediatric vaccination programmes in the early 2000s, beginning with the seven-valent pneumococcal conjugate vaccine (PCV7), led to a rapid shift in pneumococcal serotype distribution [2]. These changes were observed not only in vaccinated children but also in unvaccinated age groups, including adults, because of strong herd protection effects [3]. Over the subsequent 25 years, the successive introduction of higher-valency PCVs has continued to reshape the pneumococcal serotype landscape [4].

Following the introduction of PCV7, substantial reductions in vaccine-targeted serotypes were observed across multiple settings, including our region [5]. These declines were driven by both direct vaccine-induced protection and indirect herd effects resulting from decreased nasopharyngeal carriage in children [6]. However, the indirect protection observed in adults was more modest. This overall limited herd effect has largely been attributed to serotype replacement, particularly by serotypes with lower invasive potential [5]. Notably, in recent years, several countries have reported an unexpected re-emergence of serotypes that had previously declined after the introduction of vaccination programs, such as serotype 4 and 19F, despite high paediatric vaccine coverage [7,8].

In Catalonia, PCV7 became available through the private market in 2001 and was subsequently replaced by PCV13, which achieved vaccination coverage rates of approximately 50% [9]. In 2016, PCV13 was incorporated into the official childhood immunisation schedule, resulting in uptake rates exceeding 95% [10], and remained in use until it was replaced by PCV15 in 2023. Regarding adult immunisation, PPSV23 was traditionally recommended for routine immunisation of individuals aged 65 years and older, as well as for adults with comorbidities. In addition, a sequential vaccination strategy (PCV13+PPSV23) was recommended for adults considered to be at high risk. In 2023, these strategies were replaced by the introduction of a single-dose regimen using PCV20, which is indicated for adults aged 65 years and older, individuals with comorbidities, and other at-risk populations [11].

Serotype 14 was one of the seven serotypes targeted by PCV7. In the pre-vaccine era, three lineages accounted for the majority of invasive disease episodes caused by serotype 14 in Catalonia [12]. The first, PMEN5 (Spain^14^-5), was a multidrug-resistant lineage associated with sequence types ST17 and ST18 and classified within Global Pneumococcal Sequence Cluster 18 (GPSC18) [12]. The second most important lineage was the β-lactams and cotrimoxazole resistant PMEN3 (Spain^9V^-3; GPSC6), which was initially associated with serotype 9V. This serotype 14 lineage emerged in the 1990s through capsular switching with a serotype 9V background [13]. However, following the introduction of PCVs, the proportion of GPSC6 serotype 9V progressively declined and eventually almost disappeared. Conversely, invasive disease caused by GPSC6 serotype 14 has persisted over the years despite the introduction of PCVs, albeit at very low frequency [4,14]. A third minor lineage associated with serotype 14 was PMEN9 (England^14^-9; GPSC18), a β-lactam-susceptible clone that as a hallmark exhibited an M phenotype of macrolide resistance conferred by the presence of *mef*(A) embedded on a defective transposon (Tn*1207.1*) [15].

After several years of minimal circulation of serotype 14, we observed a marked resurgence of this serotype in Catalonia three years after the SARS-CoV-2 pandemic. Elucidating the factors driving the re-emergence of a vaccine-covered serotype as a cause of invasive pneumococcal disease (IPD) in adults is essential for anticipating future epidemiological shifts. To address this, we conducted a comprehensive investigation of serotype 14 that integrated (i) an assessment of epidemiological trends (2023–2025) across Catalonia within the framework of a laboratory-based surveillance programme, (ii) an analysis of the temporal dynamics of circulating lineages over a 30-year period at our hospital and (iii) an in-depth genomic characterisation of this serotype.

## METHODS

### Study design and clinical data

This laboratory-based study analysed serotype 14 isolates causing IPD in adults (≥18 years), with IPD defined as the isolation of *S. pneumoniae* from a normally sterile site in a patient presenting with signs and symptoms of infection. Clinical data of patients with IPD due to serotype 14 were collected, including demographics, clinical presentation, comorbidities, 30-day mortality and pneumococcal vaccination history from clinical records.

To address the three main objectives, two study collections were established. The first is a historical collection (C1) that comprised all pneumococcal isolates recovered between 1996 and 2025 at the Microbiology Department of Hospital Universitari de Bellvitge (HUB). The second is a multicentre collection that (C2) encompassed isolates from seven tertiary care hospitals across Catalonia (2023–2025), representing all four provinces: Barcelona (Hospital Universitari de Bellvitge, Hospital Universitari Germans Trias i Pujol, and Hospital Universitari Vall d’Hebron), Girona (Hospital Universitari Dr. Josep Trueta), Lleida (Hospital Universitari Arnau de Vilanova), and Tarragona (Hospital Universitari Joan XXIII and Hospital Universitari Verge de la Cinta). For genomic analysis propose, Genomic Sets (GS) were stablished including strains form C1 and C2 as well closed genomes form public databases. The overall study design and is summarised in Supplementary Figure S1.

### Serotyping, antibiotic susceptibility testing, and molecular typing

Pneumococcal isolates were identified using standard microbiological procedures and serotyped at the Spanish Pneumococcal Reference Laboratory. Antimicrobial susceptibility testing was performed by broth microdilution, with results interpreted according to the current breakpoints of the European Committee on Antimicrobial Susceptibility Testing (EUCAST) (www.eucast.org/).

Within the first collection (HUB), three distinct typing methodologies have been employed over the study period: pulsed-field gel electrophoresis (PFGE) following restriction with SmaI [16] and/or multilocus sequence typing (MLST) [17] for isolates obtained between 1996 and 2022. From 2023 onwards, all isolates from both collections have been routinely subjected to Illumina short-read whole-genome sequencing (WGS), as described in detail in the following section.

### Whole genome sequencing and bioinformatic analysis

Genomic DNA was extracted using the MagMAX™ DNA Multi-Sample Ultra 2.0 Kit on a KingFisher Flex automated system (Thermo Fisher Scientific) and quantified using a Qubit Flex Fluorometer (Thermo Fisher Scientific). Sequencing libraries were prepared using the DNA Prep Library Preparation Kit and subjected to paired-end sequencing on a MiSeq or NextSeq platform (Illumina).

Sequence quality assessment and genome assembly were performed using the Bactopia [18] pipeline, incorporating read quality control with FastQC, assembly with Shovill and annotation with Prokka. The MLST, cgMLST and the LIN codes were obtained from PubMLST. The *in silico* serotype (SeroBA), the PBP alleles (PBPtyper), the Global Pneumococcal Sequencing Cluster (GPSC) and the antibiotic resistance acquired genes and amino acid substitution were obtained from PathogenWatch (pathogen.watch/). Phylogenetic trees were reconstructed by IQ-tree [19] using the alignment obtained by Snippy (github.com/tseemann/snippy). Best model was determined by ModelFinder and 1000 UFBoot replicates were used.

For capsular operon analysis, long reads sequencing was performed in a selection of isolates. DNA was extracted using MagMAX™ DNA Multi-Sample Ultra 2.0 Kit on a KingFisher Flex automated system (Thermo Fisher Scientific) and libraries were prepared using the Native Barcoding Kit (SQK-NBD114.24) and sequenced on R10.4.1 flow cells (Oxford Nanopore Technologies). Bioinformatic analysis was conducted using Bactopia [18], which includes hybrid assembly (Unicycler). Complete capsular operon were analysed in Geneious R9 using serotype 14 (accession: CR931662) as reference. Extended bioinformatic methodology is summarized in Supplementary Table S1. Metadata for all sequenced isolates, including European Nucleotide Archive (ENA) accession numbers, are provided in Supplementary Table S2.

## RESULTS

### Historical trends and genetic lineages of serotype 14 causing IPD in adults (Hospital Universitari de Bellvitge, 1996-2025)

During the study period, 2,631 adult IPD episodes were recorded at HUB. From 1996 onwards, the annual number of IPD episodes increased progressively, reaching a peak of 147 episodes in 2009. After the introduction of PCV13, the number of IPD episodes markedly declined to 75 episodes in 2011 and remained stable with occasional fluctuations until 2019. After the emergence of the SARS-CoV-2 pandemic, IPD incidence decreased markedly, with only 30 episodes recorded in 2021, but subsequently recovered to pre-pandemic levels from 2023 onwards. Focusing on PCV7 serotypes, their proportion declined progressively from 43.4% in 1996 to 8.5% in 2019. However, a marked increase was observed in 2025, when they accounted for 17.6% of IPD episodes. Overall, serotype 14 represented 5.9% of cases (n=155), although its proportion varied substantially during the study period, declining from 13.2% in 1996 to 0.0% in 2016. The 2025 rise in PCV7 serotypes was largely driven by serotype 14, which accounted for 11.0% of all IPD episodes (Figure 1).

**Figure 1.**
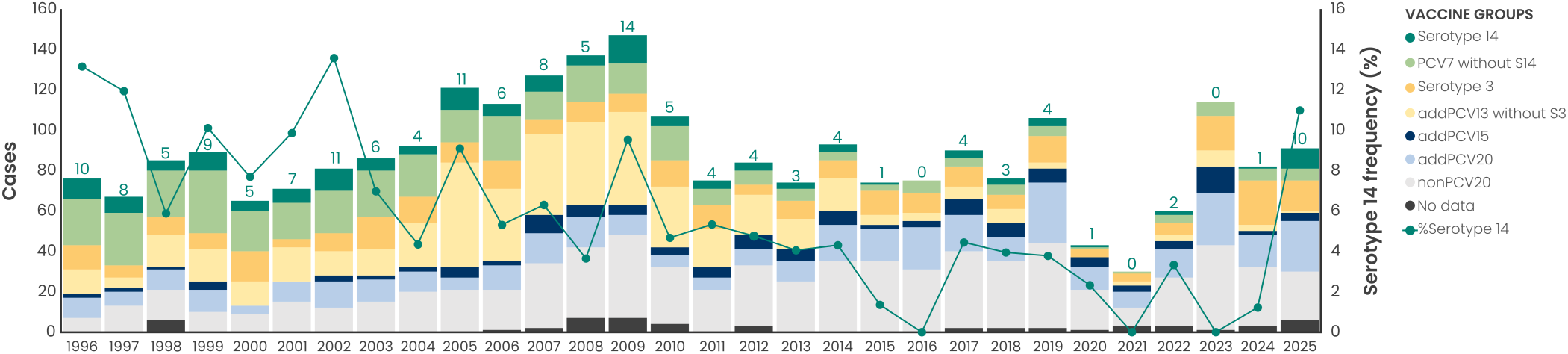
Serotypes dynamics at Hospital Universitari de Bellvitge (1996–2025). Bars represent the number of cases per year (left axis), with serotypes grouped by vaccine type: nonPCV20 serotypes (light grey), additional PCV20 serotypes (addPCV20; 8, 10A, 11A, 12F, and 15B; light blue), additional PCV15 serotypes (addPCV15; 22F and 33F; dark blue), additional PCV13 serotypes (addPCV13; 6A and 19A; yellow), and PCV7 serotypes (4, 6B, 9V, 18C, 19F, and 23F; light green). Serotype 14 (PCV7) and serotype 3 (PCV13) are highlighted independently in dark green and orange, respectively. Cases with non-available serotype are marked in dark grey. The line shows the frequency of serotype 14 over the study period (right axis).

Focusing on the genetic lineages of serotype 14, GPSC6 was the predominant lineage throughout the study period, accounting for 77.4% of isolates. Nevertheless, several other lineages were identified, including GPSC18 (13.5%), GPSC16 (1.9%), and GPSC39 (1.9%), together with additional minor lineages, highlighting the genetic heterogeneity of this serotype. GPSC18 reached its highest prevalence in 1997, exceeding that of GPSC6 (62.5% vs 25.0%), but declined, remaining at low levels until it disappeared after 2011. Notably, GPSC18 isolates were detected again in 2025, indicating the re-emergence of this historical lineage (Figure 2).

**Figure 2.**
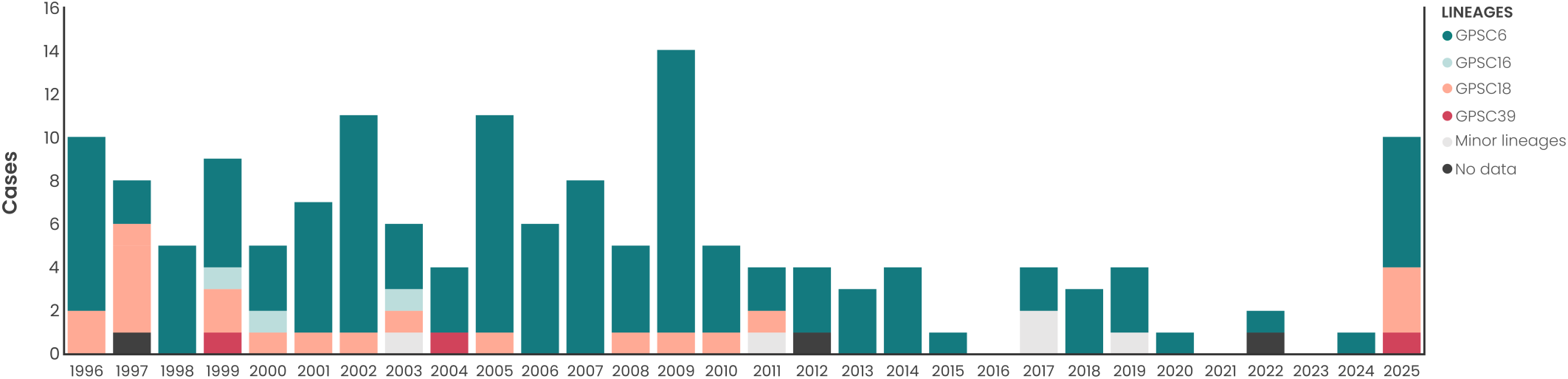
Evolution of serotype 14 lineages at Hospital Universitari de Bellvitge (1996–2025). Bars represent the number of cases per year for each genomic lineage: GPSC6 (dark green), GPSC16 (light blue), GPSC18 (salmon), GPSC39 (pink), minor lineages (light grey), and cases with lineage not available (dark grey).

### Current burden and clinical profile of serotype 14 IPD patients in Catalonia (2023-2025)

As part of the Catalan IPD surveillance programme, a total of 1,231 adult IPD episodes were collected across different geographical regions of Catalonia: 399 in 2023, 397 in 2024, and 435 in 2025. Serotype 14, as observed at HUB, increased in frequency from 0.75% (n=3) in 2023 to 2.5% (n=10) in 2024 and 10.8% (n=45) in 2025 (Figure 3A).

**Figure 3.**
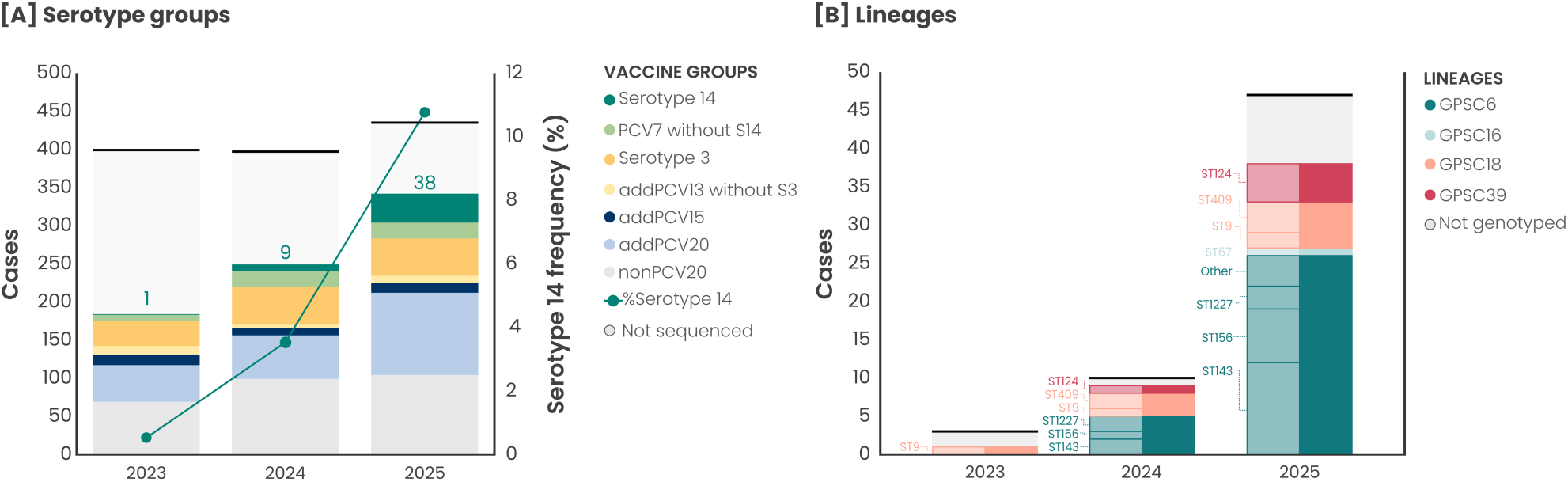
Trends of serotype 14 in Catalonia (2023-2025). **[A]** The number of cases is represented on the left axis, with serotypes grouped by vaccine type: nonPCV20 serotypes (light grey), additional PCV20 serotypes (addPCV20; 8, 10A, 11A, 12F, and 15B; light blue), additional PCV15 serotypes (addPCV15; 22F and 33F; dark blue), additional PCV13 serotypes (addPCV13; 6A and 19A; yellow), and PCV7 serotypes (4, 6B, 9V, 18C, 19F, and 23F; light green). Serotype 14 (PCV7) and serotype 3 (PCV13) are highlighted independently in dark green and orange, respectively. Cases with non-available serotype are marked in light grey. The line shows the frequency of serotype 14 over the study period (right axis). **[B]** Bars represent the number of cases per year for each genomic lineage: GPSC6 (dark green), GPSC16 (light blue), GPSC18 (salmon), GPSC39 (pink), and cases with lineage not available (light grey). Each GPSC is subdivided into different STs detected.

Among patients with IPD caused by serotype 14 with available clinical data (n=48), the mean age was 62.8 years, and 56.3% of patients were female. The main clinical presentation was pneumonia, accounting for 83.3% of cases, while meningitis represented 6.3%. Overall, 56.3% of patients presented one or more comorbidities, with malignancies being the most frequent (22.9%). The 29.2% of patients presented sepsis and 18.8% were admitted to the ICU. The 30-day mortality rate was 8.3%. Among the 48 patients, only a small proportion (18.8%) had been appropriately vaccinated. Of them, 10.4% had received PCV20, 4.2% PCV13, and 4.2% PPV23 (Table 1). Vaccinated patients were older, with a mean age of 69.6 years, and all had at least one underlying comorbidity. The most frequently reported conditions were malignancy and immunosuppressive therapy (4/9 patients each). Notably, no deaths were recorded among vaccinated patients (Supplementary Table S3).

**Table 1.** Characteristics of 48 patients with IPD due to serotype 14 in Catalonia (2023-2025).

| **Clinical characteristics** | **n** | **%** |
|---|---|---|
| **Age(years), mean(±SD)** | 62.8 (±15.8) |  |
| **Female** | 26 | 56.3 |
| **Clinical presentation** |  |  |
| Pneumonia | 40 | 83.3 |
| Meningitis | 3 | 6.3 |
| **Comorbidities** |  |  |
| One or more | 27 | 56.3 |
| Chronic pulmonary disease | 7 | 14.6 |
| Chronic heart disease | 7 | 14.6 |
| Diabetes mellitus | 10 | 20.8 |
| Malignancies | 11 | 22.9 |
| Liver cirrhosis | 2 | 4.2 |
| HIV infection | 0 | 0.0 |
| Chronic renal failure (EIV-V) | 2 | 4.2 |
| Cerebrovascular disease/dementia | 0 | 0.0 |
| Immunosuppressive therapy | 6 | 12.5 |
| **Severity and outcome** |  |  |
| Pleural effusion | 8 | 16.7 |
| Sepsis | 14 | 29.2 |
| Respiratory failure | 11 | 22.9 |
| ICU admission | 9 | 18.8 |
| 30-day mortality | 4 | 8.3 |
| **Pneumococcal vaccination** |  |  |
| Correctly vaccinated | 9 | 18.8 |
| PPV23 (less than 5 years) | 2 | 4.2 |
| PCV13 | 2 | 4.2 |
| PCV20 | 5 | 10.4 |

### Genetic lineages associated with the re-emergence of serotype 14 in Catalonia

WGS analysis identified four distinct serotype 14 lineages associated with IPD in Catalonia, all of which had previously been detected in the HUB collection. GPSC6 was the most prevalent lineage, accounting for five cases in 2024 and 26 cases in 2025. This lineage exhibited substantial genetic diversity and comprised several sequence types (STs), including ST143, ST156, ST1227, and six additional minor STs. GPSC18 accounted for one case in 2023, 3 in 2024, and 6 in 2025, all of them ST9 or ST409. All isolates of GPSC39 were ST124 (1 in 2024 and 5 in 2025) and a single case of GPSC16 (ST67) was detected in 2025 (Figure 3B).

Figure 4 displays the phylogenetic tree of the 48 sequenced serotype 14 genomes from Catalonia. GPSC6 presented the highest diversity, with many PBP profiles and differences in antibiotic resistance mechanisms. ST156 was resistant to penicillin and amoxicillin and non-susceptible to cefotaxime; in addition, the I100L substitution in DHFR conferred resistance to cotrimoxazole. The same β-lactam resistance profile was shown by ST143, which also harboured the *erm*(B) gene, conferring resistance to macrolides and clindamycin. ST1227 carried both *erm*(B) and the *aph(3’)-IIIa* gene, the latter conferring resistance to aminoglycosides. Both STs of GPSC18 were susceptible to β-lactams and shared the same PBP profile, but ST9 genomes carried the *mef*(A) gene, conferring resistance to macrolides. The single GPSC16 (ST67) strain was resistant to penicillin and amoxicillin. Strains of ST124 (GPSC39) were fully susceptible to antibiotics, without any acquired resistance genes in their genomes.

**Figure 4.**
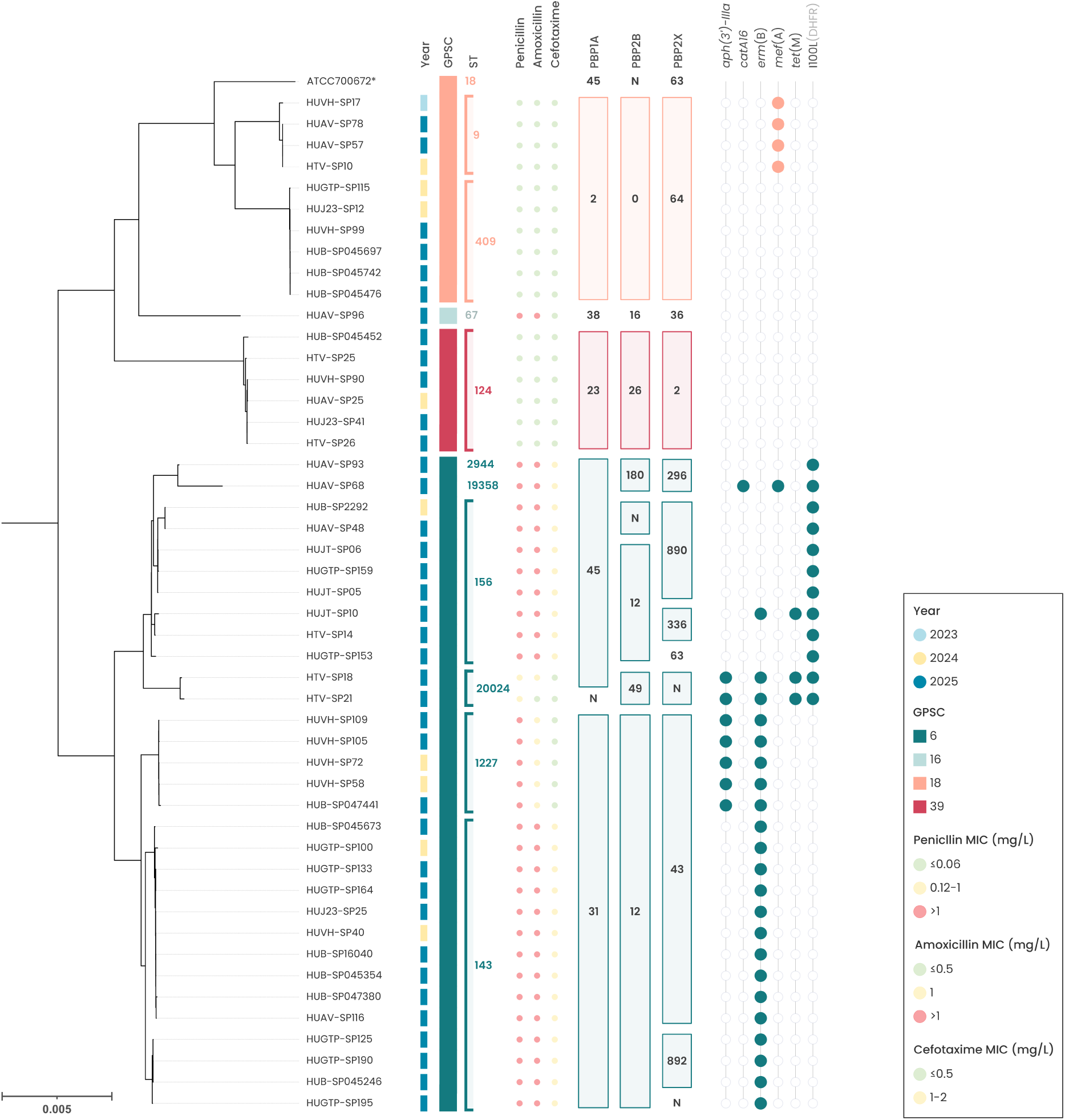
Phylogenetic tree of serotype 14 genomes from Catalonia (2023–2025). Associated metadata include year of isolation, GPSC, ST, MIC for penicillin, amoxicillin, and cefotaxime, PBP1A, 2B, and 2X alleles, acquired resistance genes, and the I100L amino acid substitution in DHFR.

To assess the genetic relatedness of the detected genomes within the broader historical serotype 14 population, an in-depth analysis was performed for each GPSC, incorporating genomes from Catalonia (GS1) and available genomes from the HUB collection (GS2). All GPSC6 genomes belonged to LIN code 0_169_0, but two main sublineages were identified: 0_169_0_10, including ST143 and ST1227, and 0_169_0_1, including ST156 and other minor lineages showing high genetic diversity (Supplementary Figure S2). GPSC9 (0_23_2) showed a new ST409 not previously detected in our setting, whereas ST17 and ST15 were only detected in the older genomes while ST9 was detected in old and new strains (Supplementary Figure S3). Regarding GPSC39 (0_61_1_1), genomes isolated in 2024–2025 showed slight genomic differences compared to those isolated in the early 2000s (Supplementary Figure S4).

### Exploring genetics of serotype 14 capsular operon

The capsular operon is the main virulence factor of pneumococcus. The capsular biosynthetic locus of serotype 14 includes 4 regulatory genes (*wzg*, *wzh*, *wzd* and *wze*), the initial transferase (*wchA*), the polymerase (*wzy*), the flippase (*wzx*) and seven glycosyl transferases (*wchJ*, *wchK*, *wchL*, *wchM*, *wchN*, *wchY* and *lrp*). The *lrp* gene encodes for a large repetitive protein that could serves as a dominant antigen [20]. The capsular operons of closely related genomes from public databases and our collection were analysed, with the aim of detecting capsular variation across the different lineages. In Supplementary Figure S5, each line represents the capsular operon of a single genome; genomes are grouped by GPSC and ST, and genes are coloured by function. Some capsular operons presented a truncated version of the *wciY* gene. The *lrp* gene contains two different repetitive sequence blocks (1 and 2). Block 1 was repeated a maximum of 18 times in HUB-SP16040, and seven capsular operons did not present any repeats. Regarding block 2, the NZ_AP028607 genome presented 14 repeats, while 7 genomes had none. There was no clear association between the number of repeats and lineage or year of isolation. Amino acid substitution of each protein are summarized in Supplementary Table S4, as well as, the number of repeats within the *lrp* gene and the length. No significant associations were identified.

## DISCUSSION

Although PCVs have substantially reduced the global burden of pneumococcal disease, their impact, particularly in adults, has been limited by the emergence and expansion of pneumococcal serotypes not included in successive PCVs [4,5]. Catalonia has long-established paediatric immunisation programme, in which PCV13 achieved an uptake above 94% between 2017 and 2022. Since 2023, children have been vaccinated with PCV15, with coverage exceeding 96%. However, the landscape of adult vaccination differs considerably. According to the official Spanish report, PPV23 uptake reached 40.1% in 2021, whereas PCV20, introduced in our region in November 2022, achieved a coverage of 26.1% by 2025 [21]. Given this limited adult coverage, the substantial decline in the incidence of PCV13 serotypes in adults, with the exception of serotype 3, is probably attributable to the indirect protection conferred by routine childhood PCV13 vaccination [22]. However, the re-emergence of vaccine serotypes has recently been reported in several countries. In England, an increase in serotype 4 has been linked to the emergence of a novel lineage, and similar trends have been observed in Spain [8,23]. Likewise, serotype 14 has re-emerged among young adults (18-59 years) in Madrid, where its incidence increased from 0.9 to 6.7 cases per 100,000 inhabitants between 2023 and 2025 [24].

In this manuscript, we describe the re-emergence of serotype 14 a cause of IPD among adults in Catalonia after the SARS-CoV-2 pandemic. Several hypotheses could explain the observed upsurge 25 years after the introduction of PCV7 into childhood vaccination. The expansion of new lineages may have contributed to this resurgence. However, our results showed that this increase was driven by multiple genetically distinct lineages, the clonal expansion of a single lineage is unlikely to fully explain this phenomenon. In fact, the rise of serotype 14 in our study was primarily associated with lineages that had been predominant in Spain prior to the introduction of PCVs [4,12]. The emergence of a capsular variant capable of evading vaccine-induced immunity may also have played a role in this upsurge. Expansion of serotype 14 following the implementation of PCV13 vaccination programmes was previously reported and attributed to the emergence of novel variants with altered capsular structures [25]. Molecular analyses have revealed diversity within the capsular operon, including truncation of *wciY* and variation in *lrp*. Our data indicate that substantial variation exists in the number of repeats within the *lrp* locus [26]. However, the biological significance of these capsular operon variations remains unclear, and their impact beyond genomic characterization has yet to be determined. Taken together, these findings argue against an exclusive role of a newly emerged vaccine-escape capsule in driving the observed expansion. Similary, the presence of a diverse genetic background suggests that the resurgence is unlikely to be driven by the acquisition of unknown non-capsular genetic determinants that selectively favoured the expansion of serotype 14 lineages.

One of the characteristics of PCVs is their ability to prevent nasopharyngeal colonisation, a prerequisite for disease development, and to induce serotype-specific memory B cells, thereby conferring long-term protection. However, weaker memory responses have been described for some serotypes, which could lead to a shorter period of protection [7]. Protection against colonisation and disease correlates with the levels of functional antibodies induced, with substantially higher antibody concentrations required to prevent colonisation than to protect against invasive disease. Compared with other pneumococcal serotypes, serotype 14 has been shown to require higher antibody concentrations to achieve protection against colonisation [27]. Furthermore, newer higher-valency conjugate vaccines have demonstrated reduced immunogenicity for serotypes shared with earlier formulations, such as PCV7 and PCV13, potentially increasing the risk of pneumococcal colonisation as predicted by modelling studies [28]. As PCV15 replaced PCV13 in the paediatric vaccination programme in Catalonia in 2023, changes in the magnitude or persistence of serotype 14-specific antibody responses could have affected the ability of the vaccine to reduce carriage of this serotype in children, potentially facilitating its transmission to adults [29]. However, we have not data on the impact of these new PCVs colonisation in children. Furthermore, serotype 14 IPD episodes occurred mostly in unvaccinated adults. The few cases identified among vaccinated individuals were largely confined to high-risk patients, including those with immunosuppression or severe underlying comorbidities who recovered after successful therapy. On the other hand, the role of adults in transmission has been increasingly recognised as important for the maintenance of most PCV serotypes [30]. With the recent changes in the epidemiology after the SARS-CoV-2 pandemic, the resurgence of different respiratory diseases makes it plausible that adults act as transmitters of vaccine serotypes. These findings suggest that the observed increase in serotype 14 incidence is unlikely to be attributable to changes in vaccination policies within the adult population.

The incidence of highly prevalent serotypes has classically been described as exhibiting fluctuating patterns [31,32], although PCV introduction has driven faster and larger shifts in serotype prevalence than those attributable to natural fluctuations [33]. Current pneumococcal serotype dynamics are likely shaped by the interplay between ecological factors, such as competition for the nasopharyngeal niche and naturally acquired population immunity, and the selective pressure exerted by vaccination [7]. In this context, it is important to note that the re-emergence of serotype 14 occurred immediately after the SARS-CoV-2 pandemic, a period that was associated with a marked global decline in the incidence of IPD due to the widespread implementation of non-pharmaceutical interventions (NPIs) [34]. Reduced interpersonal contact during the pandemic may have limited population-level exposure to pneumococcal serotypes, potentially resulting in a global decline in natural protective immunity, consistent with the concept of an immunity gap [35]. It is therefore plausible that NPIs, as well as their subsequent lifting, had differential impacts on the circulation and transmission of distinct pneumococcal serotypes. In this context, the expansion of serotype 14, which requires higher antibody levels to prevent colonisation, may have been favoured.

This study has several notable strengths. First, the collection spans a long period, allowing the analysis of serotype distribution and lineage dynamics over time. Second, an in-depth genomic analysis was performed within each lineage, allowing detailed characterisation of sequence types, antibiotic resistance determinants, and capsular locus variability, and providing valuable insight into the molecular epidemiology of serotype 14, including its genetic diversity and evolution over time. Several limitations should also be acknowledged. Although the multicentre collection included seven hospitals, it did not cover the entire Catalan region, which has approximately 8 million inhabitants, potentially limiting the generalisability of the findings. Additionally, the study focused exclusively on adults, characterising serotype 14 dynamics among children would also be highly relevant, given that transmission patterns may differ between age groups. Furthermore, clinical data were not available for patients with IPD caused by non-serotype 14 pneumococci, limiting our ability to identify potential serotype-specific differences in clinical presentation and outcomes.

In conclusion, different factors, or a combination of them, may have contributed to the re-emergence of serotype 14 in adults in our setting. Notably, vaccination coverage among adults remains low, which is particularly concerning in a population with increasing life expectancy and a growing number of comorbidities. Further studies are warranted to monitor the evolving dynamics of serotype 14 and other PCV serotypes as vaccination coverage increases.

## Data availability

Sequence data generated in this study have been deposited in the European Nucleotide Archive (ENA) under project accession numbers PRJEB24965, PRJEB76656, PRJEB80707, PRJEB93786, and PRJEB129720. Individual isolate accession numbers are listed in Supplementary Table S2.

## Ethical statement

This work was conducted in accordance with the Declaration of Helsinki of the World Medical Association and approved by the Clinical Research Ethics Committee of Hospital Universitari de Bellvitge (PR283/21). The requirement for written informed consent was waived as this study was retrospective and observational. All the data were anonymized, and patient confidentiality was always protected in accordance with current laws in Spain (LOPD3/2018 and RD1720/2007).

## Acknowledgements

We would like to thank to the staff of the Microbiology Laboratory of collaborator hospitals who contributed with their daily work to this project. We thank also the CERCA Programme/Generalitat de Catalunya for institutional support. Preliminary results of this study were presented at the 14th Meeting of the International Society of Pneumonia and Pneumococcal Disease (ISPPD) in Copenhagen, 2026.

## REFERENCE

[1] GBD 2019 Antimicrobial Resistance Collaborators. Global mortality associated with 33 bacterial pathogens in 2019: a systematic analysis for the Global Burden of Disease Study 2019. The Lancet 2022;400:2221–48. 10.1016/S0140-6736(22)02185-7.

[2] Horner C, Reynolds R, Mushtaq S, Chaudhry A, Adkin R, Allen M, et al. Trends of serotypes and resistance among *Streptococcus pneumoniae* in the UK and Ireland (1999–2019). Journal of Antimicrobial Chemotherapy 2025;80:iv72–86. 10.1093/jac/dkaf253.

[3] Shiri T, Datta S, Madan J, Tsertsvadze A, Royle P, Keeling MJ, et al. Indirect effects of childhood pneumococcal conjugate vaccination on invasive pneumococcal disease: a systematic review and meta-analysis. vol. 5. 2017. 10.1016/S2214-109X(16)30306-0.

[4] Calvo-Silveria S, González-Díaz A, Marimón JM, Cercenado E, Quesada MD, Casabella A, et al. Resilience and emergence of pneumococcal serotypes and lineages in adults post-PCV13 in Spain: A multicentre study. J Infect Public Health 2025;18. 10.1016/j.jiph.2024.102619.

[5] Ardanuy C, Tubau F, Pallares R, Calatayud L, Domínguez MA, Rolo D, et al. Epidemiology of invasive pneumococcal disease among adult patients in Barcelona before and after pediatric 7-valent pneumococcal conjugate vaccine introduction, 1997-2007. Clinical Infectious Diseases 2009;48:57–64. 10.1086/594125.

[6] Sá-Leão R, Nunes S, Brito-Avô A, Frazão N, Simões AS, Crisóstomo MI, et al. Changes in pneumococcal serotypes and antibiotypes carried by vaccinated and unvaccinated day-care centre attendees in Portugal, a country with widespread use of the seven-valent pneumococcal conjugate vaccine. Clinical Microbiology and Infection 2009;15:1002–7. 10.1111/j.1469-0691.2009.02775.x.

[7] Thindwa D, Carcamo PM, Dagan R, Weinberger DM. What is driving the resurgence and persistence of vaccine-targeted pneumococcal serotypes?—Serotype 19F as the paradigm. PLoS Pathog 2026;22. 10.1371/journal.ppat.1014065.

[8] Pérez-García C, Llorente J, Alastuey MEA, Llamosí M, Gil-Prieto R, Laghlali G, et al. Lineage dynamics and risk factors underlying serotype 4 invasive pneumococcal disease in Spain. J Infect Dis 2026. 10.1093/infdis/jiag326.

[9] Domínguez Á, Ciruela P, Hernández S, García-García JJ, Soldevila N, Izquierdo C, et al. Effectiveness of the 13-valent pneumococcal conjugate vaccine in preventing invasive pneumococcal disease in children aged 7-59 months. A matched case-control study. PLoS One 2017;12. 10.1371/journal.pone.0183191.

[10] de Sevilla MF, Alcaraz-Soler C, Soldevila N, Izquierdo C, Esteva C, Moraga-Llop F, et al. Clinical manifestations, serotype distribution, and incidence of pediatric invasive pneumococcal disease in Catalonia (Spain), 2018–2022. Eur J Pediatr 2025;184. 10.1007/s00431-025-06137-1.

[11] Manual de vacunacions i immunitzacions de Catalunya -Capítol 4. Vacunes disponibles a Catalunya. 2026.

[12] Ardanuy C, Marimón JM, Calatayud L, Giménez M, Alonso M, Grau I, et al. Epidemiology of invasive pneumococcal disease in older people in Spain (2007-2009): Implications for future vaccination strategies. PLoS One 2012;7. 10.1371/journal.pone.0043619.

[13] Càmara J, Cubero M, Martín-Galiano AJ, García E, Grau I, Nielsen JB, et al. Evolution of the b-lactam-resistant *Streptococcus pneumoniae* PMEN3 clone over a 30 year period in Barcelona, Spain. Journal of Antimicrobial Chemotherapy 2018;73:2941–51. 10.1093/jac/dky305.

[14] Pérez-García C, Sempere J, de Miguel S, Hita S, Úbeda A, Vidal EJ, et al. Surveillance of invasive pneumococcal disease in Spain exploring the impact of the COVID-19 pandemic (2019-2023). Journal of Infection 2024;89. 10.1016/j.jinf.2024.106204.

[15] Calatayud L, Ardanuy C, Cercenado E, Fenoll A, Bouza E, Pallares R, et al. Serotypes, clones, and mechanisms of resistance of erythromycin-resistant *Streptococcus pneumoniae* isolates collected in Spain. Antimicrob Agents Chemother 2007;51:3240–6. 10.1128/AAC.00157-07.

[16] Tenover FC, Arbeit RD, Goering R V, Mickelsen PA, Murray BE, Persing DH, et al. Interpreting Chromosomal DNA Restriction Patterns Produced by Pulsed-Field Gel Electrophoresis: Criteria for Bacterial Strain Typing. vol. 33. 1995. 10.1128/jcm.33.9.2233-2239.1995.

[17] Enright MC, Spratt BG. A multilocus sequence typing scheme for *Streptococcus pneumoniae*: identification of clones associated with serious invasive disease. vol. 144. n.d. 10.1099/00221287-144-11-3049.

[18] Petit RA, Read TD. Bactopia: a Flexible Pipeline for Complete Analysis of Bacterial Genomes. MSystems 2020;5. 10.1128/msystems.00190-20.

[19] Nguyen LT, Schmidt HA, Von Haeseler A, Minh BQ. IQ-TREE: A fast and effective stochastic algorithm for estimating maximum-likelihood phylogenies. Mol Biol Evol 2015;32:268–74. 10.1093/molbev/msu300.

[20] Bentley SD, Aanensen DM, Mavroidi A, Saunders D, Rabbinowitsch E, Collins M, et al. Genetic analysis of the capsular biosynthetic locus from all 90 pneumococcal serotypes. PLoS Genet 2006;2:0262–9. 10.1371/journal.pgen.0020031.

[21] Ministerio de Sanidad - Gobierno de España. SIVAMI - Informe de evolución de coberturas de vacunación por vacuna n.d. https://pestadistico.inteligenciadegestion.sanidad.gob.es/publicoSNS/I/sivamin/informe-de-evolucion-de-coberturas-de-vacunacion-por-vacuna (accessed September 1, 2026).

[22] Redin A, Ciruela P, de Sevilla MF, Gomez-Bertomeu F, Gonzalez-Peris S, Benitez MA, et al. Serotypes and Clonal Composition of Streptococcus pneumoniae Isolates Causing IPD in Children and Adults in Catalonia Systematic Introduction of PCV13. 2021.

[23] Bertran M, D’Aeth JC, Abdullahi F, Eletu S, Andrews NJ, Ramsay ME, et al. Invasive pneumococcal disease 3 years after introduction of a reduced 1 + 1 infant 13-valent pneumococcal conjugate vaccine immunisation schedule in England: a prospective national observational surveillance study. Lancet Infect Dis 2024;24:546–56. 10.1016/S1473-3099(23)00706-5.

[24] Sanz-Moreno JC, Garrido-Estepa M, Íñigo-Martínez J, Martín-Martínez F, Humanes-Navarro AM, Garrido-Buenache A, et al. Increase in serotypes 4 and 14 in invasive pneumococcal disease in adults aged 18 to 59 years. A retrospective population-based descriptive study during the period 2007–2025 in the Community of Madrid, Spain. European Journal of Clinical Microbiology and Infectious Diseases 2026. 10.1007/s10096-026-05624-z.

[25] Cave R, Kalizang’oma A, Chaguza C, Mwalukomo TS, Kamng’ona A, Brown C, et al. Expansion of pneumococcal serotype 23F and 14 lineages with genotypic changes in capsule polysaccharide locus and virulence gene profiles post introduction of pneumococcal conjugate vaccine in Blantyre, Malawi. Microb Genom 2024;10. 10.1099/mgen.0.001264.

[26] van Tonder AJ, Bray JE, Quirk SJ, Haraldsson G, Jolley KA, Maiden MCJ, et al. Putatively novel serotypes and the potential for reduced vaccine effectiveness: capsular locus diversity revealed among 5405 pneumococcal genomes. Microb Genom 2016;2:000090. 10.1099/mgen.0.000090.

[27] Voysey M, Fanshawe TR, Kelly DF, O’Brien KL, Kandasamy R, Shrestha S, et al. Serotype-Specific Correlates of Protection for Pneumococcal Carriage: An Analysis of Immunity in 19 Countries. Clinical Infectious Diseases 2018;66:913–20. 10.1093/cid/cix895.

[28] Wong A, Warren JL, Fitch L, Perniciaro S, Dagan R, Weinberger DM. Estimating the Serotype-Specific Association Between the Concentration of Vaccine-Induced Serum Antibodies and Protection Against Pneumococcal Colonisation. Journal of Infectious Diseases 2025;232:e27–34. 10.1093/infdis/jiaf106.

[29] Greenberg D, Hoover PA, Vesikari T, Peltier C, Hurley DC, McFetridge RD, et al. Safety and immunogenicity of 15-valent pneumococcal conjugate vaccine (PCV15) in healthy infants. Vaccine 2018;36:6883–91. 10.1016/j.vaccine.2018.02.113.

[30] Safadi D El, Hitchins L, Howard A, Aley P, Bowman J, Bertran M, et al. Pneumococcal Carriage and Disease in Adults in England, 2011–2019: The Importance of Adults as a Reservoir for Pneumococcus in Communities. Journal of Infectious Diseases 2025;231:e17–27. 10.1093/infdis/jiae351.

[31] Fenoll A, Granizo JJ, Aguilar L, Giménez MJ, Aragoneses-Fenoll L, Hanquet G, et al. Temporal trends of invasive *Streptococcus pneumoniae* serotypes and antimicrobial resistance patterns in Spain from 1979 to 2007. J Clin Microbiol 2009;47:1012–20. 10.1128/JCM.01454-08.

[32] Paulo AC, Sá-Leão R. Periodic cycles of pneumococcal serotypes carried by children before and after 7-valent pneumococcal conjugate vaccine. PLoS One 2017;12. 10.1371/journal.pone.0176723.

[33] Weinberger M, Weinberger DM, Malley R, Lipsitch M. Serotype replacement in disease after pneumococcal vaccination. WwwThelancetCom 2011;378:1962–73. 10.1016/S0140.

[34] Shaw D, Abad R, Amin-Chowdhury Z, Bautista A, Bennett D, Broughton K, et al. Trends in invasive bacterial diseases during the first 2 years of the COVID-19 pandemic: analyses of prospective surveillance data from 30 countries and territories in the IRIS Consortium. Lancet Digit Health 2023;5:e582–93. 10.1016/S2589-7500(23)00108-5.

[35] Cohen R, Ashman M, Taha MK, Varon E, Angoulvant F, Levy C, et al. Pediatric Infectious Disease Group (GPIP) position paper on the immune debt of the COVID-19 pandemic in childhood, how can we fill the immunity gap? Infect Dis Now 2021;51:418–23. 10.1016/j.idnow.2021.05.004.

