## Supplementary material for "Old lineages, new threat: the re-emergence of pneumococcal serotype 14 in adults in Catalonia, Spain (2023-2025)"

**Supplementary Table S1.** Extended summary of bioinformatic methodology

| PROGRAM | VERSION | STEP | REFERENCE | PARAMETERS | WEB | PAPER |
|---|---|---|---|---|---|---|
| **BACTOPIA** | **3.0.0** | **Complete analysis of bacterial genomes** | | Default | https://bactopia.github.io/latest/ | 1 |
| **BACTOPIA GATHER MODULE** | | **Get all the samples into a single place** | | Default | | |
| art | 2.5.8 | For simulating error-free reads for an input assembly | | Default | https://www.niehs.nih.gov/research/resources/software/biostatistics/art | 2 |
| fastq-scan | 1.0.1 | Reads a FASTQ from STDIN and outputs summary statistics | | Default | https://github.com/rpetit3/fastq-scan | |
| **BACTOPIA QC MODULE** | | **Quality control on Illumina reads** | | Default | | |
| fastp | 0.23.4 | A tool designed to provide fast all-in-one preprocessing for FastQ files | | Default | https://github.com/OpenGene/fastp | 3 |
| fastqc | 0.12.1 | A quality control tool for high throughput sequence data | | Default | https://www.bioinformatics.babraham.ac.uk/projects/fastqc/ | |
| lighter | 1.1.2 | A tool for correcting sequencing errors in Illumina reads | | Default | https://github.com/mourisl/Lighter | 4 |
| **BACTOPIA ASSEMBLER MODULE** | | **Assembly of Illumina reads** | | Default | | |
| any2fasta | 0.4.2 | Convert various sequence formats to FASTA | | Default | https://github.com/tseemann/any2fasta | |
| assembly-scan | 1.0.0 | Summary statistics of an input assembly | | Default | https://github.com/rpetit3/assembly-scan | |
| bwa | 0.7.17-r1188 | Mapping DNA sequences against a large reference genome | | Default | https://github.com/lh3/bwa/ | 5 |
| flash | 1.2.11 | Merge paired-end reads | | Default | https://ccb.jhu.edu/software/FLASH/ | 6 |
| megahit | 1.2.9 | Ultra-fast and memory-efficient NGS assembler | | Default | https://github.com/voutcn/megahit | 7 |
| samclip | 0.4.0 | Filter SAM file for soft and hard clipped alignments | | Default | https://github.com/tseemann/samclip | |
| samtools | '1.18' | Interacting with high-throughput sequencing data | | Default | https://github.com/samtools/samtools | 8 |
| shovill | 1.1.0 | Assemble bacterial isolate genomes from Illumina paired-end reads | | Default | https://github.com/tseemann/shovill | |
| skesa | 2.5.1 | Strategic k-mer extension for scrupulous assemblies | | Default | https://github.com/ncbi/SKESA | 9 |
| spades | 3.15.5 | Assembly and analysis of sequencing data | | Default | https://github.com/ablab/spades | 10 |
| velvet | 1.2.10 | Algorithms for de novo short read assembly | | Default | https://github.com/dzerbino/velvet | 11 |
| **BACTOPIA UNICYCLER** | | | | | | |
| medaka | 1.11.3 | Create consensus sequences and variant calls from nanopore sequencing data | | Default | https://github.com/nanoporetech/medaka | |
| miniasm | 0.3-r179 | Fast OLC-based de novo assembler for noisy long reads. | | Default | https://github.com/lh3/miniasm | 12 |
| minimap2 | 2.28-r1209 | Efficiently find multiple approximate mapping positions between two sets of long sequences | | Default | https://github.com/lh3/minimap | 12 |
| nanoq | 0.10.0 | Ultra-fast quality control and summary reports for nanopore reads | | Default | https://github.com/esteinig/nanoq | |
| pilon | '1.24' | Comprehensive microbial variant detection and genome assembly improvement | | Default | https://github.com/broadinstitute/pilon/wiki | 13 |
| racon | 1.5.0 | Fast and accurate de novo genome assembly from long uncorrected reads | | Default | https://github.com/isovic/racon | 14 |

| | | | | | | |
|---|---|---|---|---|---|---|
| rasusa | 0.8.0 | Randomly subsample sequencing reads or alignments. | | Default | https://github.com/mbhall88/rasusa | 15 |
| raven | 1.8.3 | De novo genome assembler for long uncorrected reads | | Default | https://github.com/lbcb-sci/raven | |
| unicycler | 0.5.0 | Resolving bacterial genome assemblies from short and long sequencing reads | | Default | https://github.com/rrwick/Unicycler | 16 |
| **BACTOPIA SKETCHER MODULE** | | **Create a sketch of the contigs, and query databases** | | Default | | |
| mash | 2.3 | Fast genome and metagenome distance estimation | | Default | https://github.com/marbl/Mash | 17 |
| sourmash | 4.8.8 | Quickly search, compare, and analyze genomic and metagenomic data sets | | Default | https://github.com/sourmash-bio/sourmash | 18 |
| **BACTOPIA PROKKA** | | **Annotate the contigs** | | Default | | |
| prokka | 1.14.6 | Rapid prokaryotic genome annotation | | Default | https://github.com/tseemann/prokka | 19 |
| **BACTOPIA SNIPPY IQTREE MODULE** | | **Pangenome/Phylogenetic Analysis** | | Default | | |
| snippy | 4.6.0 | Rapid haploid variant calling and core genome alignment | | Default | https://github.com/tseemann/snippy | |
| snp-dists | 0.8.2 | Convert a FASTA alignment to SNP distance matrix | | Default | https://github.com/tseemann/snp-dists | 20 |
| iqtree | 2.2.2.7 | Efficient and versatile phylogenomic software by maximum likelihood | | 1000 UFBoot replicates | https://github.com/Cibiv/IQ-TREE | |
| | | ***General tree*** | ATCC70672 | TVM+F+I+G4 | | |
| | | ***GPSC6*** | HUB-SP16040 | K3Pu+F+I | | |
| | | ***GPSC18*** | ATCC700676 | K3Pu+F+I | | |
| | | ***GPSC39*** | ATCC6314 | K3Pu+F | | |
| **PUBMLST** | **online** | **Public databases for molecular typing and microbial genome diversity** | | | https://pubmlst.org/ | 21 |
| MLST | | Multilocus sequence typing scheme scheme for *S. pneumoniae* | | | https://pubmlst.org/organisms/streptococcus-pneumoniae | 22 |
| cgMLST | | Core genome multilocus sequence typing scheme for *S. pneumoniae* | | | https://pubmlst.org/bigsdb?db=pubmlst_spneumoniae_isolates_pgl | 23 |
| LIN code | | Taxonomic life identification number barcoding system | | | https://pubmlst.org/bigsdb?db=pubmlst_spneumoniae_isolates_pgl | 23 |
| **PATHOGENWATCH** | **online** | **Platform for genomic surveillance of microbial pathogens** | | | https://pathogen.watch/ | |
| SeroBA | | In silico pneumoccal serotyping | | | | 24 |
| PBPtyper | | Predicting β-Lactam Resistance Levels in S. pneumoniae | | | | 25 |
| **OTHER** | | | | | | |
| iTol | online | Tool for the management, display, annotation and manipulation of phylogenetic and other trees | | | https://itol.embl.de/ | 26 |
| Geneious | R9 | Bioinformatic software for sequence data analysis | | CR931662 | https://www.geneious.com/updates/geneious-prime-r9-1 | |

### References

1. Petit III RA, Read TD Bactopia - a flexible pipeline for complete analysis of bacterial genomes. mSystems 5 (2020)
2. Huang W, Li L, Myers JR, Marth GT ART: a next-generation sequencing read simulator. Bioinformatics 28, 593–594 (2012)

3. Chen S, Zhou Y, Chen Y, and Gu J fastp: an ultra-fast all-in-one FASTQ preprocessor. Bioinformatics, 34(17), i884–i890. (2018)
4. Song L, Florea L, Langmead B Lighter: Fast and Memory-efficient Sequencing Error Correction without Counting. Genome Biol. 15(11):509 (2014)
5. Li H Aligning sequence reads, clone sequences and assembly contigs with BWA-MEM. arXiv [q-bio.GN] (2013)
6. Magoč T, Salzberg SL FLASH: fast length adjustment of short reads to improve genome assemblies. Bioinformatics 27.21 2957-2963 (2011)
7. Li D, Liu C-M, Luo R, Sadakane K, Lam T-W MEGAHIT: an ultra-fast single-node solution for large and complex metagenomics assembly via succinct de Bruijn graph. Bioinformatics 31.10 1674-1676 (2015)
8. Li H, Handsaker B, Wysoker A, Fennell T, Ruan J, Homer N, Marth G, Abecasis G, Durbin R The Sequence Alignment/Map format and SAMtools. Bioinformatics 25, 2078–2079 (2009)
9. Souvorov A, Agarwala R, Lipman DJ SKESA: strategic k-mer extension for scrupulous assemblies. Genome Biology 19:153 (2018)
10. Bankevich A, Nurk S, Antipov D, Gurevich AA, Dvorkin M, Kulikov AS, Lesin VM, Nikolenko SI, Pham S, Prjibelski AD, Pyshkin AV, Sirotkin AV, Vyahhi N, Tesler G, Alekseyev MA, Pevzner PA SPAdes: a new genome assembly algorithm and its applications to single-cell sequencing. Journal of computational biology 19.5 455-477 (2012)
11. Zerbino DR, Birney E Velvet: algorithms for de novo short read assembly using de Bruijn graphs. Genome research 18.5 821-829 (2008)
12. Li, Heng. “Minimap and miniasm: fast mapping and de novo assembly for noisy long sequences.” Bioinformatics (Oxford, England) vol. 32,14 (2016): 2103-10. doi:10.1093/bioinformatics/btw152
13. Walker, Bruce J et al. “Pilon: an integrated tool for comprehensive microbial variant detection and genome assembly improvement.” PloS one vol. 9,11 e112963. 19 Nov. 2014, doi:10.1371/journal.pone.0112963
14. Vaser, Robert et al. “Fast and accurate de novo genome assembly from long uncorrected reads.” Genome research vol. 27,5 (2017): 737-746. doi:10.1101/gr.214270.116
15. Hall, M. B., (2022). Rasusa: Randomly subsample sequencing reads to a specified coverage. Journal of Open Source Software, 7(69), 3941, https://doi.org/10.21105/joss.03941
16. Wick RR, Judd LM, Gorrie CL, Holt KE. Unicycler: resolving bacterial genome assemblies from short and long sequencing reads. PLoS Comput Biol 2017.
17. Ondov BD, Starrett GJ, Sappington A, Kostic A, Koren S, Buck CB, Phillippy AM Mash Screen: high-throughput sequence containment estimation for genome discovery Genome Biol 20, 232 (2019)
18. Brown CT, Irber L sourmash: a library for MinHash sketching of DNA. JOSS 1, 27 (2016)
19. Seemann T Prokka: rapid prokaryotic genome annotation Bioinformatics 30, 2068–2069 (2014)
20. Nguyen L-T, Schmidt HA, von Haeseler A, Minh BQ IQ-TREE: A fast and effective stochastic algorithm for estimating maximum likelihood phylogenies. Mol. Biol. Evol. 32:268-274 (2015)
21. Jolley KA, Bray JE, Maiden MCJ Open-access bacterial population genomics: BIGSdb software, the PubMLST.org website and their applications. Wellcome Open Res 3, 124 (2018)
22. Enright, Mark C, and Brian G Spratt. “A multilocus sequence typing scheme for Streptococcus pneumoniae: identification of clones associated with serious invasive disease.” Microbiology (Reading, England) vol. 144 ( Pt 11) (1998): 3049-3060. doi:10.1099/00221287-144-11-3049
23. Jansen van Rensburg, Melissa J et al. “Development of the Pneumococcal Genome Library, a core genome multilocus sequence typing scheme, and a taxonomic life identification number barcoding system to investigate and define pneumococcal population structure.” Microbial genomics vol. 10,8 (2024): 001280. doi:10.1099/mgen.0.001280
24. Lorenz, Oliver et al. “SeroBA(v2.0) and SeroBAnk: a robust genome-based serotyping scheme and comprehensive atlas of capsular diversity in Streptococcus pneumoniae.” Microbial genomics vol. 11,10 (2025): 001483. doi:10.1099/mgen.0.001483
25. Li, Yuan et al. “Penicillin-Binding Protein Transpeptidase Signatures for Tracking and Predicting β-Lactam Resistance Levels in Streptococcus pneumoniae.” mBio vol. 7,3 e00756-16. 14 Jun. 2016, doi:10.1128/mBio.00756-16
26. Letunic, Ivica, and Peer Bork. “Interactive Tree of Life (iTOL) v6: recent updates to the phylogenetic tree display and annotation tool.” Nucleic acids research vol. 52,W1 (2024): W78-W82. doi:10.1093/nar/gkae268

**Supplementary Table S2.** Metadata of study genomes.

| | Genome | Collection | Genomic Set | ENA Project | ENA Accession | Year | Age | Sex | Sample |
|---|---|---|---|---|---|---|---|---|---|
| 1 | HUB-SP03890 | C1 | GS2 | PRJEB24965 | ERR2303052 | 1996 | >75 | Male | Blood |
| 2 | HUAV-SP116 | C2 | GS1 | PRJEB129720 | ERR18119561 | 2025 | 18-50 | Female | Blood |
| 3 | HUB-SP045246 | C1+C2 | GS1 | PRJEB129720 | ERR18119573 | 2025 | 65-74 | Male | Blood |
| 4 | HUB-SP045354 | C1+C2 | GS1 | PRJEB129720 | ERR18119574 | 2025 | 51-64 | Female | Blood |
| 5 | HUB-SP045673 | C1+C2 | GS1 | PRJEB129720 | ERR18119577 | 2025 | 51-64 | Male | Blood |
| 6 | HUB-SP047380 | C1+C2 | GS1 | PRJEB129720 | ERR18119580 | 2025 | 18-50 | Female | Blood |
| 7 | HUB-SP06181 | C1 | GS2 | PRJEB24965 | ERR2303083 | 2001 | >75 | Female | Blood |
| 8 | HUB-SP16040 | C1+C2 | GS1+GS3 | PRJEB129720 | ERR18119593 | 2025 | 18-50 | Male | Blood |
| 9 | HUGTP-SP164 | C2 | GS1 | PRJEB129720 | ERR18157049 | 2025 | 51-64 | Female | Blood |
| 10 | HUGTP-SP190 | C2 | GS1 | PRJEB129720 | ERR18157050 | 2025 | >75 | Female | Blood |
| 11 | HUGTP-SP195 | C2 | GS1 | PRJEB129720 | ERR18157051 | 2025 | 18-50 | Male | Blood |
| 12 | HUGTP-SP125 | C2 | GS1 | PRJEB129720 | ERR18157045 | 2025 | >75 | Female | Blood |
| 13 | HUGTP-SP133 | C2 | GS1 | PRJEB129720 | ERR18157046 | 2025 | >75 | Female | Blood |
| 14 | HUGTP-SP100 | C2 | GS1 | PRJEB129720 | ERR18157043 | 2024 | 51-64 | Female | Blood |
| 15 | HUJ23-SP25 | C2 | GS1 | PRJEB129720 | ERR18119605 | 2025 | 18-50 | Male | Blood |
| 16 | HUVH-SP40 | C2 | GS1 | PRJEB129720 | ERR18157055 | 2024 | 65-74 | Female | Blood |
| 17 | HTV-SP14 | C2 | GS1 | PRJEB129720 | ERR18119556 | 2025 | 18-50 | Female | Blood |
| 18 | HUAV-SP48 | C2 | GS1 | PRJEB129720 | ERR18119563 | 2025 | 18-50 | Female | Blood |
| 19 | HUB-SP03746 | C1 | GS2+GS3 | PRJEB129720 | ERR18119569 | 1996 | 18-50 | Male | Blood |
| 20 | HUB-SP04941 | C1 | GS2 | PRJEB24965 | ERR2303077 | 1999 | 18-50 | Male | Blood |
| 21 | HUB-SP05959 | C1 | GS2 | PRJEB24965 | ERR2303096 | 2001 | 65-74 | Male | Blood |
| 22 | HUB-SP06352 | C1 | GS2 | PRJEB24965 | ERR2303078 | 2002 | 65-74 | Male | Blood |
| 23 | HUB-SP06427 | C1 | GS2 | PRJEB24965 | ERR2303084 | 2002 | >75 | Female | Blood |
| 24 | HUB-SP06646 | C1 | GS2 | PRJEB24965 | ERR2303079 | 2003 | 18-50 | Male | Blood |
| 25 | HUB-SP07116 | C1 | GS2 | PRJEB24965 | ERR2303080 | 2004 | 51-64 | Male | Blood |
| 26 | HUB-SP08170 | C1 | GS2 | PRJEB93786 | ERR15314411 | 2006 | 51-64 | Male | Cerebrospinal fluid |
| 27 | HUB-SP08293 | C1 | GS2 | PRJEB24965 | ERR2303081 | 2006 | 51-64 | Female | Blood |
| 28 | HUB-SP08416 | C1 | GS2 | PRJEB93786 | ERR15314413 | 2007 | 18-50 | Female | Cerebrospinal fluid |
| 29 | HUB-SP09671 | C1 | GS2 | PRJEB24965 | ERR2303085 | 2009 | 65-74 | Male | Blood |
| 30 | HUB-SP10356 | C1 | GS2 | PRJEB24965 | ERR2303086 | 2010 | 51-64 | Male | Blood |
| 31 | HUB-SP10568 | C1 | GS2 | PRJEB24965 | ERR2303087 | 2011 | >75 | Female | Blood |
| 32 | HUB-SP11165 | C1 | GS2 | PRJEB24965 | ERR2303088 | 2012 | 18-50 | Female | Blood |

| | | | | | | | | | |
|---|---|---|---|---|---|---|---|---|---|
| 33 | HUB-SP11208 | C1 | GS2 | PRJEB24965 | ERR2303089 | 2013 | 18-50 | Female | Blood |
| 34 | HUB-SP11617 | C1 | GS2 | PRJEB24965 | ERR2303090 | 2014 | 65-74 | Male | Blood |
| 35 | HUB-SP13538 | C1 | GS2 | PRJEB76656 | ERR13347886 | 2019 | 51-64 | Female | Blood |
| 36 | HUB-SP14172 | C1 | GS2 | PRJEB76656 | ERR13347954 | 2020 | 65-74 | Female | Blood |
| 37 | HUB-SP15283 | C1 | GS2 | PRJEB83272 | ERR14057086 | 2022 | >75 | Male | Blood |
| 38 | HUB-SP2292 | C1+C2 | GS1 | PRJEB129720 | ERR18119594 | 2024 | 65-74 | Male | Cerebrospinal fluid |
| 39 | HUGTP-SP153 | C2 | GS1 | PRJEB129720 | ERR18157047 | 2025 | 65-74 | Male | Blood |
| 40 | HUGTP-SP159 | C2 | GS1 | PRJEB129720 | ERR18157048 | 2025 | 65-74 | Male | Blood |
| 41 | HUJT-SP05 | C2 | GS1 | PRJEB129720 | ERR18119607 | 2025 | 18-50 | Female | Blood |
| 42 | HUJT-SP06 | C2 | GS1 | PRJEB129720 | ERR18119608 | 2025 | >75 | Female | Blood |
| 43 | HUJT-SP10 | C2 | GS1 | PRJEB129720 | ERR18119609 | 2025 | 65-74 | Male | Blood |
| 44 | HUB-SP047441 | C1+C2 | GS1 | PRJEB129720 | ERR18119581 | 2025 | 18-50 | Female | Blood |
| 45 | HUB-SP13722 | C1 | GS2 | PRJEB76656 | ERR13350317 | 2019 | >75 | Female | Blood |
| 46 | HUVH-SP105 | C2 | GS1 | PRJEB129720 | ERR18157052 | 2025 | 18-50 | Male | Blood |
| 47 | HUVH-SP58 | C2 | GS1 | PRJEB129720 | ERR18157056 | 2024 | 65-74 | Male | Blood |
| 48 | HUVH-SP72 | C2 | GS1 | PRJEB129720 | ERR18157057 | 2024 | >75 | Female | Blood |
| 49 | HUVH-SP109 | C2 | GS1 | PRJEB129720 | ERR18157053 | 2025 | >75 | Male | Blood |
| 50 | HUAV-SP93 | C2 | GS1 | PRJEB129720 | ERR18119567 | 2025 | 65-74 | Female | Blood |
| 51 | HUB-SP08746 | C1 | GS2 | PRJEB24965 | ERR2303092 | 2007 | >75 | Male | Blood |
| 52 | HUB-SP07744 | C1 | GS2 | PRJEB24965 | ERR2303093 | 2005 | 51-64 | Male | Blood |
| 53 | HUAV-SP68 | C2 | GS1 | PRJEB129720 | ERR18119565 | 2025 | >75 | Female | Blood |
| 54 | HTV-SP18 | C2 | GS1 | PRJEB129720 | ERR18119557 | 2025 | >75 | Female | Blood |
| 55 | HTV-SP21 | C2 | GS1 | PRJEB129720 | ERR18119558 | 2025 | >75 | Female | Blood |
| 56 | HUB-SP04519 | C1 | GS2 | PRJEB24965 | ERR2303076 | 1998 | 65-74 | Female | Blood |
| 57 | HUB-SP08540 | C1 | GS2 | PRJEB24965 | ERR2303094 | 2007 | 65-74 | Male | Transthoracic neddle aspiration |
| 58 | HUB-SP13676 | C1 | GS2+GS3 | PRJEB76656 | ERR13347900 | 2019 | >75 | Female | Blood |
| 59 | HUAV-SP96 | C2 | GS1 | PRJEB129720 | ERR18119568 | 2025 | >75 | Male | Blood |
| 60 | HUB-SP05601 | C1 | GS2+GS3 | PRJEB129720 | ERR18119583 | 2000 | 65-74 | Male | Blood |
| 61 | HTV-SP10 | C2 | GS1 | PRJEB129720 | ERR18119555 | 2024 | 51-64 | Female | Blood |
| 62 | HUAV-SP57 | C2 | GS1 | PRJEB129720 | ERR18119564 | 2025 | 51-64 | Male | Cerebrospinal fluid |
| 63 | HUAV-SP78 | C2 | GS1 | PRJEB129720 | ERR18119566 | 2025 | 51-64 | Male | Blood |
| 64 | HUB-SP06045 | C1 | GS2 | PRJEB129720 | ERR18119584 | 2001 | 18-50 | Male | Cerebrospinal fluid |
| 65 | HUB-SP09040 | C1 | GS2 | PRJEB129720 | ERR18119587 | 2008 | >75 | Female | Pleural fluid |
| 66 | HUB-SP10359 | C1 | GS2 | PRJEB129720 | ERR18119589 | 2010 | 18-50 | Male | Blood |

| | | | | | | | | | |
|---|---|---|---|---|---|---|---|---|---|
| 67 | HUVH-SP17 | C2 | GS1 | PRJEB129720 | ERR18157054 | 2023 | 65-74 | Male | Blood |
| 68 | HUB-SP04104 | C1 | GS2+GS3 | PRJEB129720 | ERR18119571 | 1997 | >75 | Male | Lung biopsy |
| 69 | HUB-SP09970 | C1 | GS2 | PRJEB129720 | ERR18119588 | 2009 | >75 | Male | Blood |
| 70 | HUB-SP10629 | C1 | GS2 | PRJEB129720 | ERR18119591 | 2011 | 18-50 | Male | Blood |
| 71 | HUB-SP04002 | C1 | GS2 | PRJEB80707 | ERR13733275 | 1996 | 18-50 | Male | Blood |
| 72 | HUB-SP04199 | C1 | GS2 | PRJEB129720 | ERR18119572 | 1997 | >75 | Male | Blood |
| 73 | HUB-SP04301 | C1 | GS2 | PRJEB80707 | ERR13733277 | 1997 | >75 | Male | Blood |
| 74 | HUB-SP04366 | C1 | GS2 | PRJEB80707 | ERR13733279 | 1997 | 18-50 | Male | Blood |
| 75 | HUB-SP04987 | C1 | GS2 | PRJEB80707 | ERR13733283 | 1999 | 18-50 | Female | Blood |
| 76 | HUB-SP06321 | C1 | GS2 | PRJEB80707 | ERR13733286 | 2002 | >75 | Female | Blood |
| 77 | HUB-SP07761 | C1 | GS2+GS3 | PRJEB80707 | ERR13733293 | 2005 | 65-74 | Male | Blood |
| 78 | HUB-SP03813 | C1 | GS2+GS3 | PRJEB129720 | ERR18119570 | 1996 | 65-74 | Male | Blood |
| 79 | HUB-SP045476 | C1+C2 | GS1 | PRJEB129720 | ERR18119576 | 2025 | >75 | Female | Blood |
| 80 | HUB-SP045697 | C1+C2 | GS1 | PRJEB129720 | ERR18119578 | 2025 | 51-64 | Male | Blood |
| 81 | HUB-SP045742 | C1+C2 | GS1+GS3 | PRJEB129720 | ERR18119579 | 2025 | 51-64 | Female | Blood |
| 82 | HUB-SP12746 | C1 | GS2 | PRJEB129720 | ERR18119592 | 2017 | 65-74 | Male | Blood |
| 83 | HUGTP-SP115 | C2 | GS1 | PRJEB129720 | ERR18157044 | 2024 | 18-50 | Male | Blood |
| 84 | HUJ23-SP12 | C2 | GS1 | PRJEB129720 | ERR18119604 | 2024 | 18-50 | Male | Blood |
| 85 | HUVH-SP99 | C2 | GS1 | PRJEB129720 | ERR18157059 | 2025 | 51-64 | Female | Blood |
| 86 | HUB-SP06948 | C1 | GS2 | PRJEB129720 | ERR18119585 | 2003 | 18-50 | Male | Blood |
| 87 | HTV-SP25 | C2 | GS1 | PRJEB129720 | ERR18119559 | 2025 | 65-74 | Male | Blood |
| 88 | HTV-SP26 | C2 | GS1 | PRJEB129720 | ERR18119560 | 2025 | 18-50 | Female | Blood |
| 89 | HUAV-SP25 | C2 | GS1 | PRJEB129720 | ERR18119562 | 2024 | 51-64 | Male | Blood |
| 90 | HUB-SP045452 | C1+C2 | GS1+GS3 | PRJEB129720 | ERR18119575 | 2025 | >75 | Female | Blood |
| 91 | HUB-SP05030 | C1 | GS2+GS3 | PRJEB129720 | ERR18119582 | 1999 | >75 | Female | Blood |
| 92 | HUB-SP07359 | C1 | GS2+GS3 | PRJEB129720 | ERR18119586 | 2004 | 65-74 | Male | Blood |
| 93 | HUJ23-SP41 | C2 | GS1 | PRJEB129720 | ERR18119606 | 2025 | 65-74 | Female | Blood |
| 94 | HUVH-SP90 | C2 | GS1 | PRJEB129720 | ERR18157058 | 2025 | 65-74 | Male | Blood |
| 95 | HUB-SP10542 | C1 | GS2+GS3 | PRJEB129720 | ERR18119590 | 2011 | >75 | Female | Blood |

| | GPSC | ST | cgST | cgST_Match | LIN | PBP1A | PBP2B | PBP2X | *aph(3')-IIIa* | *catA* | *catA16* | *erm*(B) | *mef*(A) | *tet*(M) |
|---|---|---|---|---|---|---|---|---|---|---|---|---|---|---|
| 1 | 6 | 44 | 27764 | 1191/1222 (97.5%) | 0_169_0_1 | 15 | 12 | 18 | | | | | | |
| 2 | 6 | 143 | 102225 | 1209/1222 (98.9%) | 0_169_0_10_47_0 | 31 | 12 | 43 | | | | X | | |
| 3 | 6 | 143 | 105501 | 1202/1222 (98.4%) | 0_169_0_10_49 | 31 | 12 | 892 | | | | X | | |

| | | | | | | | | | | | | | |
|---|---|---|---|---|---|---|---|---|---|---|---|---|---|
| 4 | 6 | 143 | 102225 | 1209/1222 (98.9%) | 0_169_0_10_47_0 | 31 | 12 | 43 | | | X | | |
| 5 | 6 | 143 | 102225 | 1216/1222 (99.5%) | 0_169_0_10_47_0_0 | 31 | 12 | 43 | | | X | | |
| 6 | 6 | 143 | 102225 | 1216/1222 (99.5%) | 0_169_0_10_47_0_0 | 31 | 12 | 43 | | | X | | |
| 7 | 6 | 143 | 28147 | 1212/1222 (99.2%) | 0_169_0_10_12_3 | 31 | 12 | 892 | X | | X | | X |
| 8 | 6 | 143 | 102225 | 1211/1222 (99.1%) | 0_169_0_10_47_0 | 31 | 12 | 43 | | | X | | |
| 9 | 6 | 143 | 102225 | 1218/1222 (99.7%) | 0_169_0_10_47_0_0_0 | 31 | 12 | 43 | | | X | | |
| 10 | 6 | 143 | 105501 | 1201/1222 (98.3%) | 0_169_0_10_49 | 31 | 12 | 892 | | | X | | |
| 11 | 6 | 143 | 105501 | 1212/1222 (99.2%) | 0_169_0_10_49_0 | 31 | 12 | NEW | | | X | | |
| 12 | 6 | 143 | 105501 | 1201/1222 (98.3%) | 0_169_0_10_49 | 31 | 12 | 892 | | | X | | |
| 13 | 6 | 143 | 102225 | 1217/1222 (99.6%) | 0_169_0_10_47_0_0 | 31 | 12 | 43 | | | X | | |
| 14 | 6 | 143 | 102225 | 1218/1222 (99.7%) | 0_169_0_10_47_0_0_0 | 31 | 12 | 43 | | | X | | |
| 15 | 6 | 143 | 102225 | 1205/1222 (98.6%) | 0_169_0_10_47 | 31 | 12 | 43 | | | X | | |
| 16 | 6 | 143 | 102225 | 1212/1222 (99.2%) | 0_169_0_10_47_0 | 31 | 12 | 43 | | | X | | |
| 17 | 6 | 156 | 106142 | 1214/1222 (99.3%) | 0_169_0_1_190_0_1 | 45 | 12 | 336 | | | | | |
| 18 | 6 | 156 | 84242 | 1186/1222 (97.1%) | 0_169_0_1 | 45 | NEW | 890 | | | | | |
| 19 | 6 | 156 | 31322 | 1207/1222 (98.8%) | 0_169_0_1_45_1 | 15 | 12 | 18 | | | | | |
| 20 | 6 | 156 | 31322 | 1190/1222 (97.4%) | 0_169_0_1 | 15 | 12 | 18 | | X | X | | X |
| 21 | 6 | 156 | 76148 | 1148/1222 (93.9%) | 0_169_0_1 | 45 | 12 | 63 | | | | | |
| 22 | 6 | 156 | 27847 | 1105/1222 (90.4%) | 0_169_0_1 | 15 | 12 | 296 | | | | | |
| 23 | 6 | 156 | 76148 | 1191/1222 (97.5%) | 0_169_0_1 | 45 | 12 | 63 | | | | | |
| 24 | 6 | 156 | 27857 | 1157/1222 (94.7%) | 0_169_0_1 | 15 | 12 | 296 | | | | X | |
| 25 | 6 | 156 | 33019 | 1208/1222 (98.9%) | 0_169_0_1_45_2 | 15 | 12 | 18 | | | | | |
| 26 | 6 | 156 | 95613 | 1201/1222 (98.3%) | 0_169_0_1_190 | 45 | 12 | 63 | | | | | |
| 27 | 6 | 156 | 27857 | 1153/1222 (94.4%) | 0_169_0_1 | 15 | 12 | 296 | | | | X | |
| 28 | 6 | 156 | 31322 | 1195/1222 (97.8%) | 0_169_0_1 | 15 | 12 | 18 | | | | | |
| 29 | 6 | 156 | 95613 | 1210/1222 (99.0%) | 0_169_0_1_190_0 | 45 | 12 | 336 | | | | | |
| 30 | 6 | 156 | 59630 | 1197/1222 (98.0%) | 0_169_0_1_0 | 45 | 12 | 63 | | | | | |
| 31 | 6 | 156 | 95613 | 1198/1222 (98.0%) | 0_169_0_1_190 | 45 | 12 | 63 | | | | | |
| 32 | 6 | 156 | 59630 | 1191/1222 (97.5%) | 0_169_0_1 | 45 | 12 | 63 | | | | | |
| 33 | 6 | 156 | 84242 | 1215/1222 (99.4%) | 0_169_0_1_197_0_0 | 45 | 12 | 890 | | | | | |
| 34 | 6 | 156 | 59630 | 1201/1222 (98.3%) | 0_169_0_1_0 | 45 | 12 | 63 | | | | | |
| 35 | 6 | 156 | 59630 | 1198/1222 (98.0%) | 0_169_0_1_0 | 45 | 12 | 63 | | | | | |
| 36 | 6 | 156 | 59630 | 1198/1222 (98.0%) | 0_169_0_1_0 | 45 | 12 | 336 | | | X | | X |
| 37 | 6 | 156 | 84242 | 1212/1222 (99.2%) | 0_169_0_1_197_0 | 45 | 12 | 890 | | | | | |

| | | | | | | | | | | | | | | |
|---|---|---|---|---|---|---|---|---|---|---|---|---|---|---|
| 38 | 6 | 156 | 84242 | 1186/1222 (97.1%) | 0_169_0_1 | 45 | NEW | 890 | | | | | | |
| 39 | 6 | 156 | 59630 | 1199/1222 (98.1%) | 0_169_0_1_0 | 45 | 12 | 63 | | | | | | |
| 40 | 6 | 156 | 84242 | 1211/1222 (99.1%) | 0_169_0_1_197_0 | 45 | 12 | 890 | | | | | | |
| 41 | 6 | 156 | 84242 | 1212/1222 (99.2%) | 0_169_0_1_197_0 | 45 | 12 | 890 | | | | | | |
| 42 | 6 | 156 | 84242 | 1210/1222 (99.0%) | 0_169_0_1_197_0 | 45 | 12 | 890 | | | | | | |
| 43 | 6 | 156 | 95613 | 1207/1222 (98.8%) | 0_169_0_1_190_0 | 45 | 12 | 336 | | | | X | | X |
| 44 | 6 | 1227 | 96263 | 1213/1222 (99.3%) | 0_169_0_10_45_0 | 31 | 12 | 43 | X | | | X | | |
| 45 | 6 | 1227 | 96263 | 1203/1222 (98.4%) | 0_169_0_10_45 | 31 | 12 | 43 | X | | | X | | |
| 46 | 6 | 1227 | 96263 | 1217/1222 (99.6%) | 0_169_0_10_45_0_0 | 31 | 12 | 43 | X | | | X | | |
| 47 | 6 | 1227 | 96263 | 1219/1222 (99.8%) | 0_169_0_10_45_0_0_0 | 31 | 12 | 43 | X | | | X | | |
| 48 | 6 | 1227 | 96263 | 1218/1222 (99.7%) | 0_169_0_10_45_0_0_0 | 31 | 12 | 43 | X | | | X | | |
| 49 | 6 | 1227 | 96263 | 1217/1222 (99.6%) | 0_169_0_10_45_0_0 | 31 | 12 | 43 | X | | | X | | |
| 50 | 6 | 2944 | 40155 | 1181/1222 (96.6%) | 0_169_0_1 | 45 | 180 | 296 | | | | | | |
| 51 | 6 | 2944 | 41384 | 1204/1222 (98.5%) | 0_169_0_1_286 | 45 | 180 | 296 | | | | | | |
| 52 | 6 | 6519 | 27934 | 1116/1222 (91.3%) | 0_169_0_1 | 45 | 76 | 330 | | | | | | |
| 53 | 6 | 19358 | 83884 | 1207/1222 (98.8%) | 0_169_0_1_391_0 | 45 | 180 | 296 | | | X | | X | |
| 54 | 6 | 20024 | 93821 | 1211/1222 (99.1%) | 0_169_0_1_423_0 | 45 | 49 | NEW | X | | | X | | X |
| 55 | 6 | 20024 | 93823 | 1215/1222 (99.4%) | 0_169_0_1_423_0_0 | NEW | 49 | NEW | X | | | X | | X |
| 56 | 6 | NW1 | 31322 | 1204/1222 (98.5%) | 0_169_0_1_45 | 15 | 12 | 18 | | | | | | |
| 57 | 6 | NW2 | 31322 | 810/1222 (66.3%) | 0_169_0 | 2 | 12 | 26 | | | | | | |
| 58 | 10 | 230 | 26124 | 1185/1222 (97.0%) | 0_15_1_11 | 17 | 46 | 20 | | | | X | | |
| 59 | 16 | 67 | 76780 | 1165/1222 (95.3%) | 0_15_0_11 | 38 | 16 | 36 | | | | | | |
| 60 | 16 | 67 | 76780 | 1198/1222 (98.0%) | 0_15_0_11_4 | 38 | 16 | 36 | | | | X | | X |
| 61 | 18 | 9 | 92842 | 1218/1222 (99.7%) | 0_23_2_4_1_43_0_0 | 2 | 0 | 64 | | | | | X | |
| 62 | 18 | 9 | 92842 | 1215/1222 (99.4%) | 0_23_2_4_1_43_0 | 2 | 0 | 64 | | | | | X | |
| 63 | 18 | 9 | 92842 | 1218/1222 (99.7%) | 0_23_2_4_1_43_0_0 | 2 | 0 | 64 | | | | | X | |
| 64 | 18 | 9 | 92051 | 1205/1222 (98.6%) | 0_23_2_4_1 | 2 | 0 | 64 | | | | | X | |
| 65 | 18 | 9 | 3535 | 1210/1222 (99.0%) | 0_23_2_4_1_10 | 2 | 0 | 64 | | | | | X | |
| 66 | 18 | 9 | 3535 | 1203/1222 (98.4%) | 0_23_2_4_1 | 2 | 0 | 64 | | | | | X | |
| 67 | 18 | 9 | 92051 | 1188/1222 (97.2%) | 0_23_2_4 | 2 | 0 | 64 | | | | | X | |
| 68 | 18 | 15 | 17189 | 1179/1222 (96.5%) | 0_23_2_11 | 2 | 0 | 64 | X | | | | | |
| 69 | 18 | 15 | 33118 | 1199/1222 (98.1%) | 0_23_2_12_5 | 15 | 31 | 18 | X | | | | | |
| 70 | 18 | 15 | 17189 | 1036/1222 (84.8%) | 0_23_2 | 2 | 0 | 64 | | | | | | |
| 71 | 18 | 17 | 7006 | 1176/1222 (96.2%) | 0_23_2_22 | 45 | 180 | NEW | | X | | X | | X |

| | | | | | | | | | | | | |
|---|---|---|---|---|---|---|---|---|---|---|---|---|
| 72 | 18 | 17 | 7006 | 1199/1222 (98.1%) | 0_23_2_22_0 | 45 | 180 | NEW | | X | X | X |
| 73 | 18 | 17 | 7006 | 1188/1222 (97.2%) | 0_23_2_22 | 45 | 180 | NEW | | X | X | X |
| 74 | 18 | 17 | 7006 | 1189/1222 (97.3%) | 0_23_2_22 | 45 | 180 | 63 | | X | X | X |
| 75 | 18 | 17 | 7006 | 1191/1222 (97.5%) | 0_23_2_22 | 45 | 180 | 296 | | X | X | X |
| 76 | 18 | 17 | 7006 | 1184/1222 (96.9%) | 0_23_2_22 | 45 | 180 | 296 | | X | X | X |
| 77 | 18 | 17 | 7006 | 1158/1222 (94.8%) | 0_23_2_22 | 45 | 180 | 296 | | X | X | X |
| 78 | 18 | 18 | 35425 | 1134/1222 (92.8%) | 0_23_2_22 | 45 | 614 | 63 | | X | X | X |
| 79 | 18 | 409 | 105633 | 1190/1222 (97.4%) | 0_23_2_13 | 2 | 0 | 64 | | | | |
| 80 | 18 | 409 | 105633 | 1185/1222 (97.0%) | 0_23_2_13 | 2 | 0 | 64 | | | | |
| 81 | 18 | 409 | 105633 | 1188/1222 (97.2%) | 0_23_2_13 | 2 | 0 | 64 | | | | |
| 82 | 18 | 409 | 105633 | 1181/1222 (96.6%) | 0_23_2_13 | 2 | 0 | 64 | | | | |
| 83 | 18 | 409 | 105633 | 1204/1222 (98.5%) | 0_23_2_13_4 | 2 | 0 | 64 | | | | |
| 84 | 18 | 409 | 105633 | 1189/1222 (97.3%) | 0_23_2_13 | 2 | 0 | 64 | | | | |
| 85 | 18 | 409 | 105633 | 1188/1222 (97.2%) | 0_23_2_13 | 2 | 0 | 64 | | | | |
| 86 | 18 | 1964 | 95542 | 1212/1222 (99.2%) | 0_23_2_12_2_1 | 15 | 31 | NEW | X | | | |
| 87 | 39 | 124 | 59513 | 1215/1222 (99.4%) | 0_61_1_1_30_20_2 | 23 | 26 | 2 | | | | |
| 88 | 39 | 124 | 59513 | 1214/1222 (99.3%) | 0_61_1_1_30_20_2 | 23 | 26 | 2 | | | | |
| 89 | 39 | 124 | 91355 | 1215/1222 (99.4%) | 0_61_1_1_30_20_2 | 23 | 26 | 2 | | | | |
| 90 | 39 | 124 | 71259 | 1208/1222 (98.9%) | 0_61_1_1_30_21 | 23 | 26 | 2 | | | | |
| 91 | 39 | 124 | 17954 | 1194/1222 (97.7%) | 0_61_1_1 | 23 | 26 | 2 | | | | |
| 92 | 39 | 124 | 17954 | 1193/1222 (97.6%) | 0_61_1_1 | 23 | 26 | 2 | | | | |
| 93 | 39 | 124 | 59513 | 1217/1222 (99.6%) | 0_61_1_1_30_20_2 | 23 | 26 | 2 | | | | |
| 94 | 39 | 124 | 91355 | 1216/1222 (99.5%) | 0_61_1_1_30_20_2 | 23 | 26 | 2 | | | | |
| 95 | 904 | 15321 | 34134 | 1151/1222 (94.2%) | 0_74_0_1 | 24 | 27 | 135 | | | | X |

**Supplementary Table S3.** Clinical characteristic of vaccinated patients

| **Clinical characteristics** | **n (%)** |
|---|---|
| Age(years), mean(±SD) | 69.6 (±14.7) |
| Female | 55.6% |
| **Clinical presentation** | |
| Pneumonia | 7 (77.8%) |
| Other | 2 (22.2%) |
| **Comorbidities** | |
| One or more | 9 (100%) |
| Chronic pulmonary disease | 3 (33.3%) |
| Chronic heart disease | 2 (22.2%) |
| Diabetes mellitus | 3 (33.3%) |
| Malignancies | 4 (44.4%) |
| Liver cirrhosis | 0 (0.0%) |
| HIV infection | 0 (0.0%) |
| Chronic renal failure (EIV-V) | 1 (11.1%) |
| Cerebrovascular disease/dementia | 0 (0.0%) |
| Immunosuppressive therapy | 4 (44.4%) |
| **Severity and outcome** | |
| Pleural effusion | 0 (0.0%) |
| Sepsis | 3 (33.3%) |
| Respiratory failure | 1 (11.1%) |
| ICU admission | 0 (0.0%) |
| 30-day mortality | 9 (100%) |

**Supplementary Table S4.** Amino acid substitution of serotype 14 capsular operon of the study genomes from Genome Collection 3 and Genomes Collection 4.

| | | | | | Wzg | | | | | | | | | | | | | | | | | | | | | | | | | | | | | | | |
|---|---|---|---|---|---|---|---|---|---|---|---|---|---|---|---|---|---|---|---|---|---|---|---|---|---|---|---|---|---|---|---|---|---|---|---|---|
| | **GENOME** | **GSPC** | **ST** | **YEAR** | 9 | 29 | 80 | 106 | 114 | 123 | 131 | 153 | 162 | 169 | 178 | 196 | 223 | 321 | 331 | 335 | 337 | 338 | 359 | 386 | 387 | 394 | 400 | 402 | 403 | 404 | 409 | 415 | 420 | 424 | 431 | 445 |
| 1 | ***CR931662*** | | | | **R** | **V** | **V** | **T** | **I** | **D** | **T** | **S** | **D** | **A** | **E** | **Y** | **N** | **M** | **H** | **E** | **S** | **A** | **G** | **L** | **Q** | **A** | **T** | **I** | **D** | **S** | **I** | **L** | **N** | **A** | **N** | **T** |
| 2 | HUB-SP16040 | 6 | 143 | 2025 | . | . | . | . | . | . | . | . | . | . | . | . | . | . | . | . | . | . | . | . | . | . | M | . | . | . | . | . | . | . | . | . |
| 3 | HUB-SP03746 | 6 | 156 | 1996 | G | . | L | A | . | E | . | . | N | . | . | . | . | L | . | . | . | . | . | . | . | . | M | . | . | . | . | . | . | . | . | . |
| 4 | HUB-SP13676 | 10 | 230 | 2019 | G | . | . | . | L | E | . | L | . | V | . | . | . | . | . | . | . | . | S | . | . | . | . | . | . | . | . | . | D | . | T | M |
| 5 | GPSC14_ref | 14 | 242 | ND | . | . | . | . | . | . | . | . | . | . | . | . | . | . | . | . | . | . | . | . | . | . | . | . | . | . | . | . | . | . | . | . |
| 6 | NZ_LR216043 | 14 | 242 | ND | . | . | . | . | . | . | . | . | . | . | . | . | . | . | . | . | . | . | . | . | . | . | . | . | . | . | . | . | . | . | . | . |
| 7 | NC_012466 | 16 | 66 | ND | . | . | . | . | . | . | . | . | . | . | . | . | . | . | . | . | . | . | . | . | . | . | M | . | . | . | . | . | . | . | . | . |
| 8 | HUB-SP05601 | 16 | 67 | 2000 | . | . | . | . | . | . | . | . | . | . | . | . | . | . | . | . | . | . | . | . | . | . | M | . | . | . | . | . | . | . | . | . |
| 9 | NZ_CP131712 | 16 | NEW | ND | . | . | . | . | . | . | . | . | . | . | . | . | . | . | . | . | . | . | . | . | . | . | . | . | . | . | . | . | . | . | . | . |
| 10 | ATCC700676 | 18 | 9 | ND | . | . | . | . | . | . | . | . | . | . | . | . | . | . | . | . | . | . | . | . | . | . | M | . | . | . | . | . | . | . | . | . |
| 11 | GPSC18_ref | 18 | 9 | ND | . | . | . | . | . | . | . | . | . | . | . | . | . | . | . | . | . | . | . | . | . | . | M | . | . | . | . | . | . | . | . | . |
| 12 | NC_017593 | 18 | 9 | ND | . | . | . | . | . | . | . | . | . | . | . | . | . | * | - | - | - | - | - | - | - | - | - | - | - | - | - | - | - | - | - | - |
| 13 | NZ_CP035263 | 18 | 9 | ND | . | . | . | . | . | . | . | . | . | . | . | . | . | . | . | . | . | . | . | . | . | . | M | . | . | . | . | . | . | . | . | . |
| 14 | NZ_LR216065 | 18 | 13 | ND | . | . | . | . | . | . | . | . | . | . | . | . | . | . | . | . | . | . | . | . | . | . | M | . | . | . | . | . | . | . | . | . |
| 15 | HUB-SP04104 | 18 | 15 | 1997 | . | . | . | . | . | . | . | . | . | . | . | . | . | . | . | . | L | . | . | . | . | . | M | . | . | . | . | . | . | . | . | . |
| 16 | NC_010582 | 18 | 15 | ND | . | I | L | . | . | . | . | . | . | . | . | . | . | . | . | . | L | . | . | . | . | . | M | . | . | . | . | . | . | . | . | . |
| 17 | NZ_LR129841 | 18 | 15 | ND | . | . | . | . | . | . | . | . | . | . | . | . | . | . | . | . | . | . | . | . | . | . | M | . | . | . | . | . | . | . | . | . |
| 18 | NZ_LR595848 | 18 | 15 | ND | . | . | . | . | . | . | . | . | . | . | . | . | . | . | . | . | . | . | . | . | . | . | M | . | . | . | . | . | . | . | . | . |
| 19 | HUB-SP07761 | 18 | 17 | 2005 | . | . | . | . | . | . | . | . | . | . | . | . | . | . | . | . | . | . | . | . | . | . | . | . | . | . | . | . | . | . | . | . |
| 20 | HUB-SP03813 | 18 | 18 | 1996 | . | . | . | . | . | . | . | . | . | . | . | . | . | . | . | . | . | . | . | . | . | . | . | . | . | . | . | . | . | . | . | . |
| 21 | ATCC700672 | 18 | 18 | ND | . | . | . | . | . | . | . | . | . | . | . | . | . | . | . | . | . | . | . | . | . | . | . | . | . | . | . | . | . | . | . | . |
| 22 | HUB-SP045742 | 18 | 409 | 2025 | . | . | . | . | . | . | . | . | . | . | . | . | . | . | . | . | L | . | . | . | . | . | M | . | . | . | . | . | . | . | . | . |
| 23 | NZ_CP035262 | 18 | 409 | ND | . | . | . | . | . | . | . | . | . | . | . | . | . | . | . | . | L | . | . | . | . | . | M | . | . | . | . | . | . | . | . | . |
| 24 | HUB-SP05030 | 39 | 124 | 1999 | . | . | . | . | . | . | . | . | . | . | . | . | . | . | . | . | . | . | . | . | . | . | . | . | . | . | . | . | . | . | . | . |
| 25 | HUB-SP07359 | 39 | 124 | 2004 | . | . | . | . | . | . | . | . | . | . | . | . | . | . | . | . | . | . | . | . | . | . | . | . | . | . | . | . | . | . | . | . |
| 26 | HUB-SP045452 | 39 | 124 | 2025 | . | . | . | . | . | . | . | . | . | . | . | . | . | . | . | . | . | . | . | . | . | . | . | . | . | . | . | . | . | . | . | . |
| 27 | ATCC6314 | 39 | 124 | ND | . | . | . | . | . | . | . | . | . | . | . | . | . | . | . | . | . | . | . | . | . | . | . | . | . | . | . | . | . | . | . | . |

| | | | | | | | | | | | | | | | | | | | | | | | | | | | | | | | | | | | | |
|---|---|---|---|---|---|---|---|---|---|---|---|---|---|---|---|---|---|---|---|---|---|---|---|---|---|---|---|---|---|---|---|---|---|---|---|---|
| 28 | GPSC39_ref | 39 | 124 | ND | . | . | . | . | . | . | . | . | . | . | . | . | . | . | . | . | . | . | . | . | . | . | . | . | . | . | . | . | . | . | . | . |
| 29 | NZ_CP035258 | 39 | 124 | ND | . | . | . | . | . | . | . | . | . | . | . | . | . | . | . | . | . | . | . | . | . | . | . | . | . | . | . | . | . | . | . | . |
| 30 | NZ_CP035259 | 39 | 124 | ND | . | . | . | . | . | . | . | . | . | . | . | . | . | . | . | . | . | . | . | . | . | . | . | . | . | . | . | . | . | . | . | . |
| 31 | NZ_LR216025 | 39 | 124 | ND | . | . | . | . | . | . | . | . | . | . | . | . | . | . | . | . | . | . | . | . | . | . | . | . | . | . | . | . | . | . | . | . |
| 32 | NZ_LS483417 | 39 | 124 | ND | . | . | . | . | . | . | . | . | . | . | . | . | . | . | . | . | . | . | . | . | . | . | . | . | . | . | . | . | . | . | . | . |
| 33 | GPSC108_ref | 108 | 9487 | ND | . | . | . | . | . | . | . | . | . | . | . | . | . | . | . | . | . | . | . | . | . | . | . | . | . | . | . | . | . | . | . | . |
| 34 | NZ_LR216028 | 108 | 9487 | ND | . | . | . | . | . | . | . | . | . | . | . | . | . | . | . | . | . | . | . | . | . | . | . | . | . | . | . | . | . | . | . | . |
| 35 | NZ_AP028607 | 279 | 343 | ND | . | . | . | . | . | E | M | . | N | . | . | H | D | L | Y | D | T | S | . | . | . | . | . | . | . | . | . | I | . | . | . | . |
| 36 | NZ_CP113114 | 904 | 7645 | ND | . | . | . | . | . | . | . | . | . | . | . | . | . | . | . | . | . | . | . | . | . | . | . | . | . | . | . | . | . | . | . | . |
| 37 | NZ_CP113115 | 904 | 7645 | ND | . | . | . | . | . | . | . | . | . | . | . | . | . | . | . | . | . | . | . | . | . | . | . | . | . | . | . | . | . | . | . | . |
| 38 | HUB-SP10542 | 904 | 15321 | 2011 | S | . | . | . | . | E | . | . | . | . | K | . | . | . | . | . | . | T | . | I | K | V | S | L | Q | G | L | I | D | T | . | M |

| | | Wzh | | | | Wzd | | | | | Wze | | | | | | | | | | | WchA | | | | | | | | | |
|---|---|---|---|---|---|---|---|---|---|---|---|---|---|---|---|---|---|---|---|---|---|---|---|---|---|---|---|---|---|---|---|
| | **GENOME** | 27 | 29 | 149 | 232 | 76 | 90 | 188 | 189 | 227 | 12 | 30 | 54 | 58 | 62 | 76 | 77 | 79 | 134 | 159 | 195 | 43 | 126 | 128 | 159 | 227 | 248 | 289 | 300 | 323 | 402 |
| 1 | ***CR931662*** | **I** | **S** | **H** | **D** | **S** | **D** | **V** | **S** | **S** | **E** | **L** | **V** | **R** | **R** | **N** | **S** | **I** | **N** | **V** | **Q** | **L** | **F** | **P** | **K** | **A** | **T** | **R** | **T** | **V** | **G** |
| 2 | HUB-SP16040 | . | . | R | . | . | . | . | . | . | . | . | . | . | . | . | . | . | . | . | . | . | . | . | . | . | . | . | . | A | . |
| 3 | HUB-SP03746 | . | . | R | . | . | . | . | . | . | . | . | . | . | . | . | . | . | . | . | . | . | . | . | . | . | . | . | . | A | . |
| 4 | HUB-SP13676 | A | . | R | . | . | . | . | . | . | . | . | . | . | . | . | . | . | . | . | . | . | . | . | . | . | . | . | . | A | . |
| 5 | GPSC14_ref | . | . | . | Y | . | . | . | . | . | . | . | . | . | . | . | . | . | . | . | . | . | . | . | . | . | . | . | . | . | . |
| 6 | NZ_LR216043 | . | . | . | Y | . | . | . | . | . | . | . | . | . | . | . | . | . | . | . | . | . | . | . | . | . | . | . | . | . | . |
| 7 | NC_012466 | . | . | R | . | . | . | . | . | . | . | . | . | . | . | . | . | . | . | . | . | . | . | . | . | . | . | . | . | A | . |
| 8 | HUB-SP05601 | . | . | R | . | . | . | . | . | . | . | . | . | . | . | . | . | . | . | . | . | . | . | . | . | . | . | . | A | A | . |
| 9 | NZ_CP131712 | . | . | . | . | L | . | . | . | . | . | . | . | . | . | . | P | . | . | . | . | . | . | . | . | . | . | . | . | . | . |
| 10 | ATCC700676 | . | . | R | . | . | . | . | . | . | . | . | . | . | . | . | . | . | . | . | . | . | . | . | . | . | . | . | . | A | . |
| 11 | GPSC18_ref | . | . | R | . | . | . | . | . | . | . | . | . | . | . | . | . | . | . | . | . | . | . | . | . | . | . | . | . | A | . |
| 12 | NC_017593 | . | . | R | . | . | . | . | . | . | . | . | . | . | . | . | . | . | . | . | . | . | . | . | . | . | . | . | . | A | . |
| 13 | NZ_CP035263 | . | . | R | . | . | . | . | . | . | . | . | . | . | . | . | . | . | . | . | . | . | . | . | E | . | . | . | . | A | A |
| 14 | NZ_LR216065 | . | . | R | . | . | . | . | . | . | . | . | . | . | . | . | . | . | . | . | . | . | . | . | . | . | . | . | . | A | . |
| 15 | HUB-SP04104 | . | . | R | . | . | . | . | . | . | . | . | . | . | . | . | . | . | . | . | . | . | . | . | . | . | . | . | . | A | . |
| 16 | NC_010582 | . | . | R | . | . | . | . | . | . | . | . | . | . | . | . | . | . | . | . | . | . | . | . | . | . | . | . | . | A | . |

| | | | | | | | | | | | | | | | | | | | | | | | | | | | | | | | |
|---|---|---|---|---|---|---|---|---|---|---|---|---|---|---|---|---|---|---|---|---|---|---|---|---|---|---|---|---|---|---|---|
| 17 | NZ_LR129841 | . | . | R | . | . | . | . | . | . | . | . | . | . | . | . | . | . | . | . | . | . | . | . | . | . | . | . | . | A | . |
| 18 | NZ_LR595848 | . | . | R | . | . | . | . | . | . | . | . | . | . | . | . | . | . | . | . | . | P | . | . | . | P | . | . | . | A | . |
| 19 | HUB-SP07761 | . | . | . | . | . | . | M | . | . | . | . | . | . | . | . | . | . | . | . | . | . | . | . | . | . | . | . | . | A | . |
| 20 | HUB-SP03813 | . | . | . | . | . | . | M | . | . | . | . | . | . | . | . | . | . | . | . | . | . | . | . | . | . | . | . | . | A | . |
| 21 | ATCC700672 | . | . | . | . | . | . | M | . | . | . | . | . | . | . | . | . | . | . | . | . | . | . | . | . | . | . | . | . | A | . |
| 22 | HUB-SP045742 | . | . | R | . | . | . | . | . | . | . | . | . | . | . | . | . | . | . | . | . | . | . | . | . | . | . | . | . | A | . |
| 23 | NZ_CP035262 | . | . | R | . | . | . | . | . | . | . | . | . | . | . | . | . | . | . | . | . | . | . | . | . | . | . | . | . | A | . |
| 24 | HUB-SP05030 | . | . | . | . | . | . | . | . | . | . | . | . | . | . | . | . | . | . | . | . | . | . | . | . | . | . | . | . | . | . |
| 25 | HUB-SP07359 | . | . | . | . | . | . | . | . | . | . | . | . | . | . | . | . | . | . | . | . | . | . | . | . | . | . | . | . | . | . |
| 26 | HUB-SP045452 | . | . | . | . | . | . | . | . | . | . | . | . | . | . | . | . | . | . | . | . | . | . | . | . | . | . | . | . | . | . |
| 27 | ATCC6314 | . | . | . | . | . | Y | . | . | . | . | . | . | . | . | . | . | . | . | . | . | . | . | . | . | . | . | . | . | . | . |
| 28 | GPSC39_ref | . | . | . | . | . | . | . | . | . | . | . | . | . | . | . | . | . | . | . | . | . | . | . | . | . | . | . | . | . | . |
| 29 | NZ_CP035258 | . | . | . | . | . | . | . | . | . | . | . | . | . | . | . | . | . | . | . | . | . | . | . | . | . | . | . | . | . | . |
| 30 | NZ_CP035259 | . | . | . | . | . | . | . | . | . | . | . | . | . | . | . | . | . | . | . | . | . | . | . | . | . | . | C | . | . | . |
| 31 | NZ_LR216025 | . | . | . | . | . | . | . | . | . | . | . | . | . | . | . | . | . | . | . | . | . | . | . | . | . | . | . | . | . | . |
| 32 | NZ_LS483417 | . | . | . | . | . | Y | . | . | . | . | . | . | . | . | . | . | . | . | . | . | . | . | . | . | . | . | . | . | . | . |
| 33 | GPSC108_ref | . | . | R | . | . | . | . | . | . | . | . | . | . | . | . | . | . | K | . | . | . | . | . | . | . | . | . | . | A | . |
| 34 | NZ_LR216028 | . | . | R | . | . | . | . | . | . | . | . | . | . | . | . | . | . | K | . | . | . | . | . | . | . | . | . | . | A | . |
| 35 | NZ_AP028607 | . | . | R | . | . | . | . | V | G | K | . | I | W | S | Q | . | M | . | I | K | . | . | T | . | . | . | . | . | A | . |
| 36 | NZ_CP113114 | . | . | R | . | . | . | . | . | . | . | F | . | . | . | . | . | . | . | . | . | . | L | . | . | . | A | . | . | A | . |
| 37 | NZ_CP113115 | . | . | R | . | . | . | . | . | . | . | F | . | . | . | . | . | . | . | . | . | . | L | . | . | . | A | . | . | A | . |
| 38 | HUB-SP10542 | A | A | R | . | . | . | . | . | . | . | . | . | . | . | . | . | . | . | . | . | . | . | . | . | . | . | . | . | A | . |

| | | WchK | | Wzy | | | WchL | | WchM | | | | | | WchN | | | | | | Wzx | | | | |
|---|---|---|---|---|---|---|---|---|---|---|---|---|---|---|---|---|---|---|---|---|---|---|---|---|---|
| | **GENOME** | 11 | 52 | 64 | 83 | 104 | 184 | 233 | 17 | 91 | 105 | 107 | 111 | 139 | 50 | 152 | 208 | 221 | 231 | 275 | 148 | 165 | 337 | 432 | 444 |
| 1 | ***CR931662*** | **Q** | **L** | **I** | **L** | **L** | **F** | **D** | **Y** | **G** | **H** | **L** | **N** | **V** | **W** | **Y** | **T** | **L** | **L** | **S** | **A** | **S** | **V** | **A** | **V** |
| 2 | HUB-SP16040 | . | V | . | I | . | . | . | . | . | . | . | . | . | . | S | I | . | F | F | T | . | . | V | . |
| 3 | HUB-SP03746 | . | V | . | I | . | . | . | . | . | . | . | . | . | . | S | I | . | F | F | T | . | . | V | . |
| 4 | HUB-SP13676 | R | . | . | I | . | . | . | . | S | S | I | D | . | . | S | I | P | . | . | T | . | . | V | I |

| | | | | | | | | | | | | | | | | | | | | | | | | | |
|---|---|---|---|---|---|---|---|---|---|---|---|---|---|---|---|---|---|---|---|---|---|---|---|---|---|
| 5 | GPSC14_ref | . | . | . | . | . | . | . | . | . | . | . | . | . | . | . | . | . | . | . | . | . | . | . | . |
| 6 | NZ_LR216043 | . | . | . | . | . | . | . | . | . | . | . | . | . | . | . | . | . | . | . | . | . | . | . | . |
| 7 | NC_012466 | . | V | . | I | . | . | . | . | . | . | . | . | . | . | S | I | . | F | F | T | . | F | V | . |
| 8 | HUB-SP05601 | . | V | . | I | . | . | . | . | . | . | . | . | . | . | S | I | . | F | F | T | N | . | V | . |
| 9 | NZ_CP131712 | . | . | . | . | . | . | . | . | . | . | . | . | . | . | . | . | . | . | . | . | . | . | . | . |
| 10 | ATCC700676 | . | V | . | I | . | . | . | . | . | . | . | . | . | C | S | I | . | F | F | T | . | . | V | . |
| 11 | GPSC18_ref | . | V | . | I | . | . | . | . | . | . | . | . | . | . | S | I | . | F | F | T | . | . | V | . |
| 12 | NC_017593 | . | V | . | I | . | . | . | . | . | . | . | . | . | . | S | I | . | F | F | T | . | . | V | . |
| 13 | NZ_CP035263 | . | V | . | I | . | . | . | . | . | . | . | . | . | . | S | I | . | F | F | T | . | . | V | . |
| 14 | NZ_LR216065 | . | V | . | I | . | . | . | . | . | . | . | . | . | . | S | I | . | F | F | T | . | . | V | . |
| 15 | HUB-SP04104 | . | V | . | I | . | L | N | . | . | . | . | . | . | . | S | I | . | F | F | T | . | . | V | . |
| 16 | NC_010582 | . | V | . | I | . | . | . | . | . | . | . | . | . | . | S | I | . | F | F | T | . | . | V | . |
| 17 | NZ_LR129841 | . | V | . | I | . | . | . | . | . | . | . | . | . | . | S | I | . | F | F | T | . | . | V | . |
| 18 | NZ_LR595848 | . | V | . | I | . | . | . | . | . | . | . | . | . | . | S | I | . | F | F | T | . | . | V | . |
| 19 | HUB-SP07761 | . | V | . | I | M | . | . | . | . | . | . | . | . | . | S | I | . | F | F | T | . | . | V | . |
| 20 | HUB-SP03813 | . | V | . | I | . | . | . | . | . | . | . | . | . | . | S | I | . | F | F | T | . | . | V | . |
| 21 | ATCC700672 | . | V | . | I | . | . | . | D | . | . | . | . | . | . | S | I | . | F | F | T | . | . | V | . |
| 22 | HUB-SP045742 | . | V | . | I | . | . | . | . | . | . | . | . | . | . | S | I | . | F | F | T | . | . | V | . |
| 23 | NZ_CP035262 | . | V | . | I | . | . | . | . | . | . | . | . | . | . | S | I | . | F | F | T | . | . | V | . |
| 24 | HUB-SP05030 | . | . | M | . | . | . | . | . | . | . | . | . | . | . | . | . | . | . | . | . | . | . | . | . |
| 25 | HUB-SP07359 | . | . | M | . | . | . | . | . | . | . | . | . | . | . | . | . | . | . | . | . | . | . | . | . |
| 26 | HUB-SP045452 | . | . | . | . | . | . | . | . | . | . | . | . | . | . | . | . | . | . | . | . | . | . | . | . |
| 27 | ATCC6314 | . | . | . | . | . | . | . | . | . | . | . | . | . | . | . | . | . | . | . | . | . | . | . | . |
| 28 | GPSC39_ref | . | . | . | . | . | . | . | . | . | . | . | . | . | . | . | . | . | . | . | . | . | . | . | . |
| 29 | NZ_CP035258 | . | . | . | . | . | . | . | . | . | . | . | . | . | . | . | . | . | . | . | . | . | . | . | . |
| 30 | NZ_CP035259 | . | . | . | . | . | . | . | . | . | . | . | . | . | . | . | . | . | . | . | . | . | . | . | . |
| 31 | NZ_LR216025 | . | . | . | . | . | . | . | . | . | . | . | . | . | . | . | . | . | . | . | . | . | . | . | . |
| 32 | NZ_LS483417 | . | . | . | . | . | . | . | . | . | . | . | . | . | . | . | . | . | . | . | . | . | . | . | . |
| 33 | GPSC108_ref | . | . | . | I | . | . | . | . | S | S | I | D | . | . | S | I | P | . | . | T | . | . | V | I |
| 34 | NZ_LR216028 | . | . | . | I | . | . | . | . | S | S | I | D | . | . | S | I | P | . | . | T | . | . | V | I |

| | | | | | | | | | | | | | | | | | | | | | | | | | |
|---|---|---|---|---|---|---|---|---|---|---|---|---|---|---|---|---|---|---|---|---|---|---|---|---|---|
| 35 | NZ_AP028607 | . | . | . | . | . | . | . | . | . | . | . | I | . | . | . | . | . | . | . | . | . | V | . |
| 36 | NZ_CP113114 | . | . | . | I | . | . | . | . | S | S | I | D | . | . | S | I | P | . | . | T | . | . | V | I |
| 37 | NZ_CP113115 | . | . | . | I | . | . | . | . | S | S | I | D | . | . | S | I | P | . | . | T | . | . | V | I |
| 38 | HUB-SP10542 | . | . | . | I | . | . | . | . | S | S | I | D | . | . | S | I | P | . | . | T | . | . | V | I |

| | | WciY | | | | | | | | | | | | | | | | | | | | | | | | | | | | | | Lrp | | | | |
|---|---|---|---|---|---|---|---|---|---|---|---|---|---|---|---|---|---|---|---|---|---|---|---|---|---|---|---|---|---|---|---|---|---|---|---|---|
| | **GENOME** | 18 | 20 | 21 | 22 | 23 | 24 | 25 | 26 | 27 | 28 | 29 | 30 | 31 | 32 | 33 | 34 | 35 | 36 | 37 | 38 | 39 | 40 | 104 | 105 | 106 | 107 | 108 | 109 | 110 | 111 | 121 | **ΔΔ LENGHT** | **B1-A** | **B1-B** | **B2-A** | **B2-B** |
| 1 | ***CR931662*** | **Y** | **V** | **V** | **T** | **N** | **K** | **Y** | **W** | **L** | **P** | **L** | **N** | **K** | **R** | **K** | **V** | **V** | **F** | **D** | **N** | **F** | **G** | **R** | **N** | **S** | **D** | **R** | **P** | **W** | **K** | **W** | 1359 | 4 | 1 | 1 | 1 |
| 2 | HUB-SP16040 | . | . | . | . | . | . | . | . | . | . | . | . | . | . | . | . | . | . | . | . | . | . | I | T | E | K | L | K | Y | * | - | 3554 | 17 | 1 | 4 | 1 |
| 3 | HUB-SP03746 | . | . | . | . | . | . | . | . | . | . | . | . | . | . | . | . | . | . | . | . | . | . | I | T | E | K | L | K | Y | * | - | 1869 | 6 | 1 | 3 | 1 |
| 4 | HUB-SP13676 | * | - | - | - | - | - | - | - | - | - | - | - | - | - | - | - | - | - | - | - | - | - | - | - | - | - | - | - | - | - | - | 2282 | 12* | 1* | 0 | 1* |
| 5 | GPSC14_ref | . | . | . | . | . | . | . | . | . | . | . | . | . | . | . | . | . | . | . | . | . | . | . | . | . | . | . | . | . | . | . | 416 | 0 | 0 | 0 | 0 |
| 6 | NZ_LR216043 | . | . | . | . | . | . | . | . | . | . | . | . | . | . | . | . | . | . | . | . | . | . | . | . | . | . | . | . | . | . | . | 416 | 0 | 0 | 0 | 0 |
| 7 | NC_012466 | . | . | . | . | . | . | . | . | . | . | . | . | . | . | . | . | . | . | . | . | . | . | I | T | E | K | L | K | Y | * | - | 2012 | 7 | 1 | 2 | 1 |
| 8 | HUB-SP05601 | . | . | . | . | . | . | . | . | . | . | . | . | . | . | . | . | . | . | . | . | . | . | I | T | E | K | L | K | Y | * | - | 1757 | 6 | 1 | 2 | 1 |
| 9 | NZ_CP131712 | . | . | . | . | . | . | . | . | . | . | . | . | . | . | . | . | . | . | . | . | . | . | . | . | . | . | . | . | . | . | . | 1235 | 0 | 1 | 5 | 1 |
| 10 | ATCC700676 | . | . | . | . | . | . | . | . | . | . | . | . | . | . | . | . | . | . | . | . | . | . | I | T | E | K | L | K | Y | * | - | 2727 | 12 | 1 | 3 | 1 |
| 11 | GPSC18_ref | . | . | . | . | . | . | . | . | . | . | . | . | . | . | . | . | . | . | . | . | . | . | I | T | E | K | L | K | Y | * | - | 1072 | 3 | 0 | 1 | 1 |
| 12 | NC_017593 | . | . | . | . | . | . | . | . | . | . | . | . | . | . | . | . | . | . | . | . | . | . | I | T | E | K | L | K | Y | * | - | 1328 | 3 | 1 | 2 | 1 |
| 13 | NZ_CP035263 | . | . | . | . | . | . | . | . | . | . | . | . | . | . | . | . | . | . | . | . | . | . | I | T | E | K | L | K | Y | * | - | 2982 | 13 | 1 | 4 | 1 |
| 14 | NZ_LR216065 | . | . | . | . | . | . | . | . | . | . | . | . | . | . | . | . | . | . | . | . | . | . | I | T | E | K | L | K | Y | * | - | 1072 | 3 | 0 | 1 | 1 |
| 15 | HUB-SP04104 | . | I | T | L | V | V | E | D | M | E | T | I | Q | S | I | L | L | M | N | Y | * | - | - | - | - | - | - | - | - | - | - | 2901 | 14 | 1 | 2 | 1 |
| 16 | NC_010582 | . | I | T | L | V | V | E | D | M | E | T | I | Q | S | I | L | L | M | N | Y | V | * | - | - | - | - | - | - | - | - | - | 787 | 0 | 1 | 1 | 1 |
| 17 | NZ_LR129841 | . | . | . | . | . | . | . | . | . | . | . | . | . | . | . | . | . | . | . | . | . | . | I | T | E | K | L | K | Y | * | - | 2951 | 12 | 1 | 5 | 1 |
| 18 | NZ_LR595848 | . | . | . | . | . | . | . | . | . | . | . | . | . | . | . | . | . | . | . | . | . | . | I | T | E | K | L | K | Y | * | - | 3013 | 14 | 1 | 3 | 1 |
| 19 | HUB-SP07761 | . | . | . | . | . | . | . | . | . | . | . | . | . | . | . | . | . | . | . | . | . | . | I | T | E | K | L | K | Y | * | - | 1695 | 4 | 1 | 4 | 1 |
| 20 | HUB-SP03813 | . | . | . | . | . | . | . | . | . | . | . | . | . | . | . | . | . | . | . | . | . | . | I | T | E | K | L | K | Y | * | - | 1726 | 5 | 1 | 3 | 1 |
| 21 | ATCC700672 | . | . | . | . | . | . | . | . | . | . | . | . | . | . | . | . | . | . | . | . | . | . | I | T | E | K | L | K | Y | * | - | 1583 | 4 | 1 | 3 | 1 |
| 22 | HUB-SP045742 | . | I | T | L | V | V | E | D | M | E | T | I | Q | S | I | L | L | M | N | Y | V | * | - | - | - | - | - | - | - | - | - | 385 | 0 | 0 | 0 | 0 |
| 23 | NZ_CP035262 | . | I | T | L | V | V | E | D | M | E | T | I | Q | S | I | L | L | M | N | Y | V | * | - | - | - | - | - | - | - | - | - | 2441 | 10 | 1 | 3 | 1 |

| | | | | | | | | | | | | | | | | | | | | | | | | | | | | | | | | | | | | | |
|---|---|---|---|---|---|---|---|---|---|---|---|---|---|---|---|---|---|---|---|---|---|---|---|---|---|---|---|---|---|---|---|---|---|---|---|---|---|
| 24 | HUB-SP05030 | . | . | . | . | . | . | . | . | . | . | . | . | . | . | . | . | . | . | . | . | . | . | . | . | . | . | . | . | . | . | . | 2584 | 11 | 1 | 3 | 1 |
| 25 | HUB-SP07359 | . | . | . | . | . | . | . | . | . | . | . | . | . | . | . | . | . | . | . | . | . | . | . | . | . | . | . | . | . | . | . | 2043 | 8 | 1 | 2 | 1 |
| 26 | HUB-SP045452 | . | . | . | . | . | . | . | . | . | . | . | . | . | . | . | . | . | . | . | . | . | . | . | . | . | . | . | . | . | . | * | 1154 | 1 | 1 | 3 | 1 |
| 27 | ATCC6314 | . | . | . | . | . | . | . | . | . | . | . | . | . | . | . | . | . | . | . | . | . | . | . | . | . | . | . | . | . | . | . | 1154 | 1 | 1 | 3 | 1 |
| 28 | GPSC39_ref | . | . | . | . | . | . | . | . | . | . | . | . | . | . | . | . | . | . | . | . | . | . | . | . | . | . | . | . | . | . | . | 416 | 0 | 0 | 0 | 0 |
| 29 | NZ_CP035258 | . | . | . | . | . | . | . | . | . | . | . | . | . | . | . | . | . | . | . | . | . | . | . | . | . | . | . | . | . | . | . | 3156 | 15 | 1 | 3 | 1 |
| 30 | NZ_CP035259 | . | . | . | . | . | . | . | . | . | . | . | . | . | . | . | . | . | . | . | . | . | . | . | . | . | . | . | . | . | . | . | 1409 | 2 | 1 | 4 | 1 |
| 31 | NZ_LR216025 | . | . | . | . | . | . | . | . | . | . | . | . | . | . | . | . | . | . | . | . | . | . | . | . | . | . | . | . | . | . | . | 416 | 0 | 0 | 0 | 0 |
| 32 | NZ_LS483417 | . | . | . | . | . | . | . | . | . | . | . | . | . | . | . | . | . | . | . | . | . | . | . | . | . | . | . | . | . | . | . | 1440 | 3 | 1 | 3 | 1 |
| 33 | GPSC108_ref | * | - | - | - | - | - | - | - | - | - | - | - | - | - | - | - | - | - | - | - | - | - | - | - | - | - | - | - | - | - | - | 437 | 1+ | 0 | 0 | 0 |
| 34 | NZ_LR216028 | * | - | - | - | - | - | - | - | - | - | - | - | - | - | - | - | - | - | - | - | - | - | - | - | - | - | - | - | - | - | - | 437 | 1+ | 0 | 0 | 0 |
| 35 | NZ_AP028607 | . | . | . | . | . | . | . | . | . | . | . | . | . | . | . | . | . | . | . | . | . | . | . | . | . | . | . | . | . | . | . | 2274 | 1 | 1 | 13 | 1 |
| 36 | NZ_CP113114 | * | - | - | - | - | - | - | - | - | - | - | - | - | - | - | - | - | - | - | - | - | - | - | - | - | - | - | - | - | - | - | 2730 | 12+ | 1+ | 4+ | 1+ |
| 37 | NZ_CP113115 | * | - | - | - | - | - | - | - | - | - | - | - | - | - | - | - | - | - | - | - | - | - | - | - | - | - | - | - | - | - | - | 2587 | 11+ | 1+ | 4+ | 1+ |
| 38 | HUB-SP10542 | * | - | - | - | - | - | - | - | - | - | - | - | - | - | - | - | - | - | - | - | - | - | - | - | - | - | - | - | - | - | - | 3016 | 12+ | 1+ | 4+ | 1+ |

ND= No data available

N+=Sequenced identity 90-99%

*lrp* gene:

- ΔΔ LENGHT = Amino acid length
- Block 1A (B1-A):

  AAGAAAAACCAACAAAACCATCAGGTAAGCTTGCGACAACTGTAGAAGTTGACGGAACAAAAGCTGATGCACAGAAAGAGCTTGAGTTGTCAGTTGCAACAGATAAAGTTACGAAGACAGTAAAA GATACTGTTGTGTACGAAAATCTGTTAGCTGGCGAAACCTACAAACTAACAGGTCAGTTG
  ATGAAGATTACAGCTGACAAAGAAGAGGAAGTAGCAACGAAAGAAACAACTTTCGTCGCAGATGCCTCAGG CAACGGTACAACCAGCCTAGAGTTTGAAGATGTGAGCCTTGAAGCAGGCGTTAAGTATGTAGTATATGAAACTGCAGAGAGCGAAAAAGAAATTGACTTCAAAGAAGGCAAAG
  AAAAGCACAAGGTAGAACACAAAGACAAAGATGACAAAGCTCAGACTGTAGTTGTTACG
- Block 1B (B1-B):

  AAAGAAAAACCAACAAAACCATCAGGTAAGCTTGCGACAACTGTAGAAGTTGACGGAACAAAAGCTGATGCACAGAAAGAGCTTGAGTTGTCAGTTGCAACAGATAAAGTTACGAAGACAGTAAAAGATACTGTTGTGTACGAAAATCTGTTAGCTGGCGAAACCTACAAACTAACAGGTCAGTT
  GATGAAGATTACAGCTGACAAAGAAGAGGAAGTAGCAACGAAAGAAACAACTTTCGTCGCAGATGCCTCAGGCAACGGTACAACCAGCCTAGAGTTTGAAGATGTGAGCCTTGAAGCAGGCGTTA AGTATGTAGTATATGAAACTGCAGAGAGCGAAAAAGAAATTGACTTCAAAGAAGGCAAA
  GAAAAGCACAAGGTAGAACACAAAGACAAAGATGACAAAGCTCAGACTGTAGTTGTATCTAAAATTAAGCCA
- Block 2A ((B2-A):

  GAGCCAGGAGCACAAGAAGTTCACTTTAGTAAAGTCAATGTAGGCGGAGAAGAAATTGCAGGTGCGGAAATTCACATCAAGCAAGGCGACACTGTCGTGGCAAGCTGGGTATCAGAAGCCGGTAA GACACATACATTGAAACTAAAACCAGGTCACTATATCTTCCATGAAGCAGTAGCTCCAGG
  TGGCTATCTAGCAGTAACAGATATTCACTTTAGTGTAGATGAAACTGGTCAAGTGACAGTAACTGATGTAAATGGAAATACAGCTGTAGCGGAAGGCAACAAGCTTACGGTTACAGACCAAACCAAACCAGTAACCCCACCAAGCCCAGAA
- Block 2B (B2-B):

  GAGCCAGGAGCACAAGAAGTTCACTTTAGTAAAGTCAATGTAGGCGGAGAAGAAATTGCAGGTGCGGAAATTCACATCAAGCAAGGCGACACTGTCGTGGCAAGCTGGGTATCAGAAGCCGGTAA GACACATACATTGAAACTAAAACCAGGTCACTATATCTTCCATGAAGCAGTAGCTCCAGG
  TGGCTATCTAGCAGTAACAGATATTCACTTTAGTGTAGATGAAACTGGTCAAGTGACAGTAACTGATGTAAATGGAAATACAGCTGTAGCGGAAGGCAACAAGCTTACGGTTACAGACCAAAGTG CTGATAAAGATAAGCAAGATAAGTTG

**Regulatory genes** **Initial trasnferase** **Polymerase** **Glycosyl trasnferase** **Flipasse**

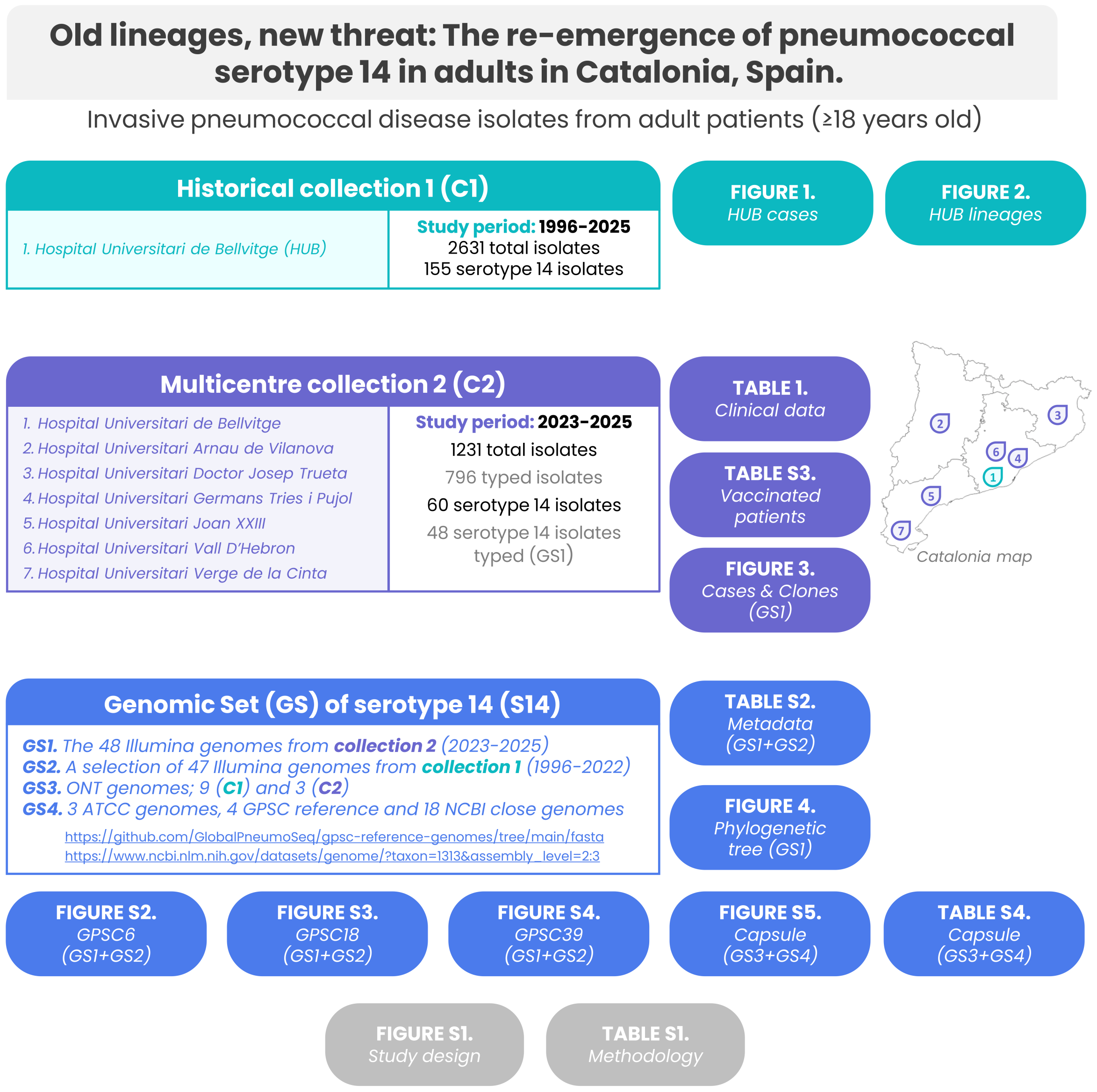

**Supplementary Figure S1. Study design of isolate collections and genomic datasets.** Collection 1 (C1, green) includes isolates from Hospital Universitari de Bellvitge collected between 1996 and 2025. Collection 2 (C2, purple) includes isolates from seven Catalan hospitals collected between 2023 and 2025. Four genomic datasets (GS1–GS4, blue) were compiled for genomic analysis purposes. In grey, figures and tables related to methodology section.

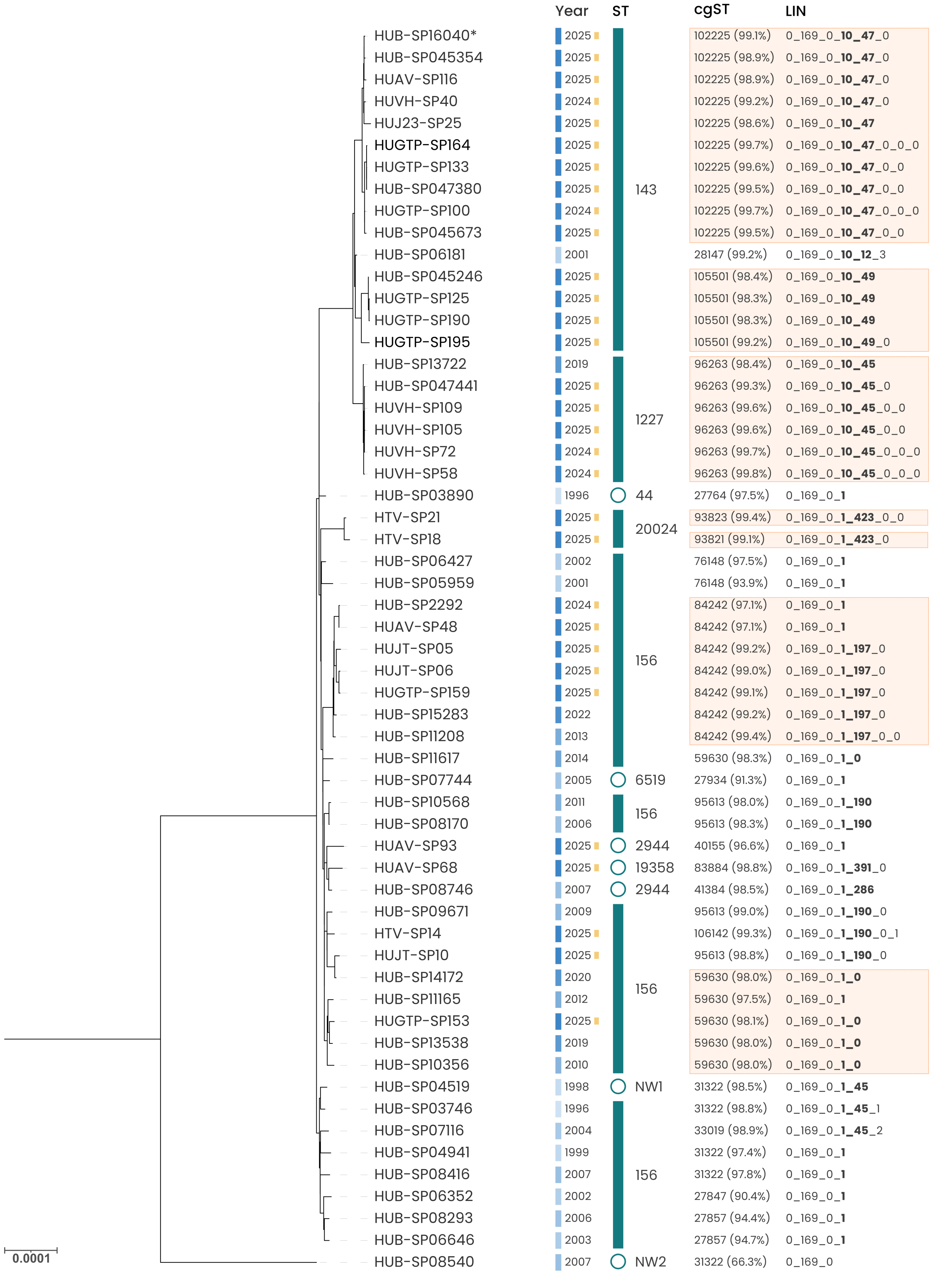

**Supplementary Figure S2. Phylogenetic tree of GPSC6 from GS1 and GS2.** Associated metadata include the year of isolation, with a yellow rectangle indicating genomes from 2023–2025, and the ST. The closest cgST and the percentage of matched alleles are indicated; cgSTs shared by more than 3 genomes are marked in orange. LIN codes are specified in the last column.

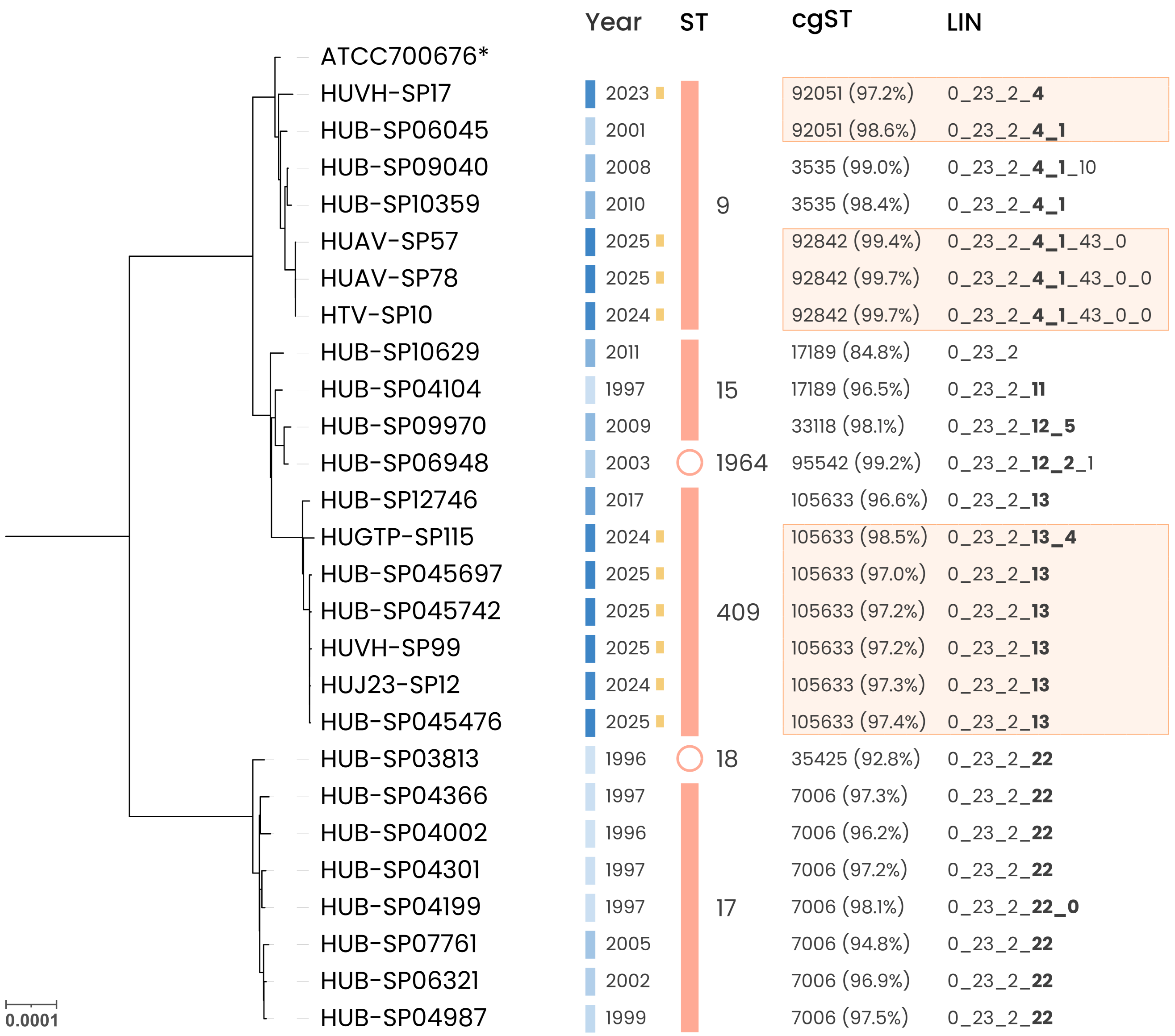

**Supplementary Figure S3. Phylogenetic tree of GPSC9 from GS1 and GS2.** Associated metadata include the year of isolation, with a yellow rectangle indicating genomes from 2023–2025, and the ST. The closest cgST and the percentage of matched alleles are indicated; cgSTs shared by more than 3 genomes are marked in orange. LIN codes are specified in the last column

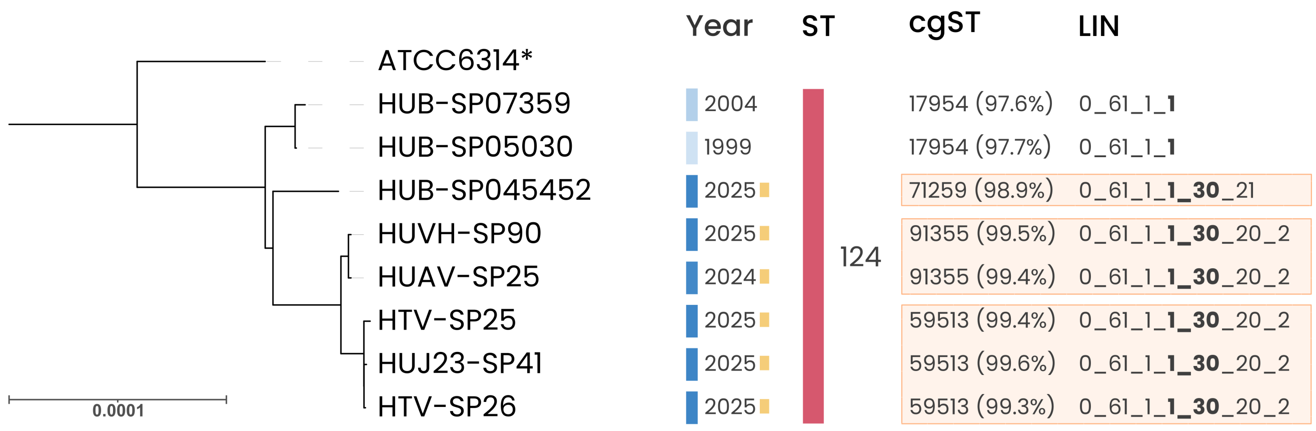

**Supplementary Figure S4. Phylogenetic tree of GPSC39 from GS1 and GS2.** Associated metadata include the year of isolation, with a yellow rectangle indicating genomes from 2023–2025, and the ST. The closest cgST and the percentage of matched alleles are indicated; cgSTs shared by more than 3 genomes are marked in orange. LIN codes are specified in the last column.

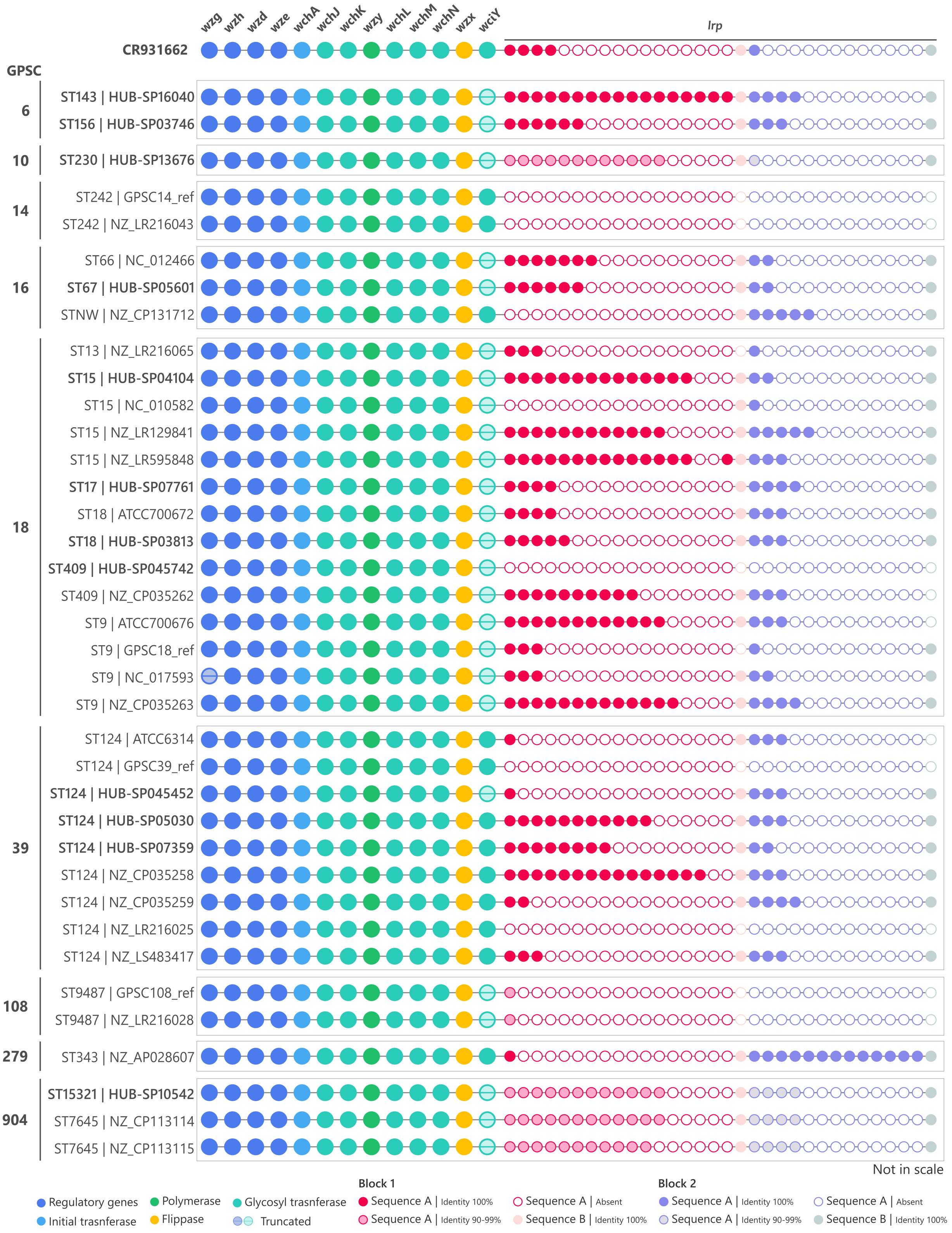

**Supplementary Figure S5. Schematic representation of the capsular operons of serotype 14 genomes.** Each line represents a capsular operon; genomes are sorted by GPSC, and the ST is indicated at the beginning of the name. Capsular genes are colored by function; regulatory genes (dark blue), initial transferase (light blue), glucosyltransferase (turquoise), polymerase (green), and flippase (yellow). Transparency in the circle fill indicates that the gene is truncated. The *lpr* gene is composed of repeats of two different blocks: block 1 (red palette) and block 2 (purple palette). Inside each block, two different sequences (A and B) can be found. Sequence B may be present or absent at the end of the block and, when present, always occurs as a single copy without repetition. When sequence A is present, it is represented by a solid circle, and when absent, by an empty circle. The repeat unit of GPSC904 shows 90–99% identity with the reference capsule sequence CR931662. The figure does not represent the actual genetic length of the genes.
